# The Efficacy and Safety of Pembrolizumab, Ipilimumab, and Nivolumab Monoteraphy and Combination for Colorectal Cancer: A Systematic Review and Meta-Analysis

**DOI:** 10.1101/2024.07.02.24309865

**Authors:** Albertus Ari Adrianto, Ignatius Riwanto, Udadi Sadhana, Dewi Kartikawati Paramita, Henry Setyawan, Kevin Christian Tjandra, Danendra Rakha Putra Respati, Derren David Christian Homenta Rampengan, Roy Novri Ramadhan, Gastin Gabriel Jangkang, Endang Mahati

## Abstract

**Background:** Colorectal cancer (CRC) ranks third globally in cancer-related mortality, with rising incidence, particularly in Asia, projecting a 60% surge by 2030. Metastatic CRC (mCRC) presents a significant challenge with a grim 5-year survival rate of 14%. Emerging evidence suggests that tumors with DNA mismatch repair deficiency (dMMR) and high microsatellite instability (MSI-H) respond well to immune checkpoint inhibitors (ICIs), marking a paradigm shift in therapeutic approaches. This systematic review and meta-analysis aim to comprehensively assess Pembrolizumab, Nivolumab, and the combination of Nivolumab and Ipilimumab in advanced CRC, considering their significant antitumor efficacy in MSI-H/dMMR mCRC.

**Methods:** Following PRISMA guidelines and Cochrane Handbook standards, this study covers 2014 to 2024, involving advanced CRC patients treated with ICIs. A comprehensive literature search employed 12 independent authors across eight databases. Parameters such as overall survival, progression-free survival, and objective response rate were extracted.The Cochrane Collaboration’s Risk of Bias version 2 tool assessed risk. Statistical analysis utilized mean difference and risk ratios with random-effect models due to anticipated heterogeneity. Robustness was ensured through publication bias analysis and sensitivity meta-analysis. Linear regression explored associations in subgroup analysis.

**Results:** The meta-analysis evaluated ORR and OS across different immunotherapy interventions. Nivolumab, Nivolumab+Ipilimumab, and Pembrolizumab exhibited varying ORR and OS effect sizes with corresponding heterogeneity levels. Progression-free survival (PFS) analysis also showed diverse effect sizes and heterogeneity levels across the three interventions. The study provides a comprehensive overview of response rates and survival outcomes for these immunotherapies in advanced CRC.

**Conclusions:** The study concludes that combination immunotherapy, particularly Nivolumab and Ipilimumab, presents a promising avenue for advanced CRC treatment, showing superior efficacy. Pembrolizumab monotherapy also exhibited promise. While the study offers valuable insights, the identified heterogeneity emphasizes the need for additional research. Adverse effects were generally low, supporting the viability of the studied immunotherapies. The study acknowledges limitations and calls for ongoing investigation to refine and validate these findings, marking a pioneering effort in systematically comparing short-term and long-term effects of anti-CTLA-4 and anti-PD-1 therapies in CRC.

## Introduction

Colorectal cancer (CRC) ranks as the third most common cause of cancer-related mortality globally[1]. Over the past thirty years, the incidence of CRC has surged in many nations, particularly in Asia, where a notable rise has been observed[2]. Projections indicate a significant rise in its worldwide incidence, with an expected increase of 60% by the year 2030 [1,3]. The prognosis remains poor for individuals diagnosed with metastatic colorectal cancer (mCRC), as evidenced by a 5-year survival rate of only 14%[4]. The incidence of CRC and its associated mortality rate rise with advancing age, with a median age of diagnosis typically reported as 68 years old. A striking 93% of CRC-related deaths are observed in individuals aged 50 years and older. The relative survival rates at 5 and 10 years post-diagnosis for CRC stand at 65% and 58%, respectively[5].

Until recently, the treatment choices available for individuals with inoperable colorectal cancer worldwide have been predominantly confined to chemotherapy and targeted therapies[6]. Findings from several clinical trials indicate that CRC tumors identified by DNA mismatch repair deficiency (dMMR) and a high level of microsatellite instability (MSI-H) exhibit susceptibility to treatment with immune checkpoint inhibitors (ICIs)[7–9]. Tumors characterized as MSI-H/dMMR possess an elevated tumor mutational burden, leading to the generation of neoantigens, thereby rendering MSI-H/dMMR tumors more immunogenic compared to those with proficient DNA mismatch repair mechanisms. Consequently, MSI-H/dMMR tumors tend to harbor increased numbers of tumor-infiltrating lymphocytes, whose function can be potentiated by ICI therapy. Patients with MSI-H/dMMR mCRC typically derive less benefit from standard chemotherapy compared to those with microsatellite stable/mismatch repair-proficient mCRC[10,11]. However, MSI-H/dMMR status serves as a predictive indicator of favorable response to anti-programmed death (PD)-1 checkpoint inhibitor therapy [8,12,13]

The interaction involving the PD-1 receptor and its ligands, PD-L1 and PD-L2, typically serves as an immune checkpoint mechanism responsible for regulating the equilibrium between T-cell activation, immune tolerance, and preventing immune-related tissue damage. This pathway is often exploited by tumors to evade immune surveillance[14]. PD-1 is present on T, B, and natural killer T cells, as well as activated monocytes and a significant portion of tumor-infiltrating lymphocytes (TILs) in various malignancies[14]. When PD-L1 (found on cells of diverse lineages) or PD-L2 (present on macrophages and dendritic cells) binds to the PD-1 receptor, it ultimately leads to the inhibition of T-cell function. Both PD-1 ligands have the potential to be expressed either constitutively or induced in various cell types, which also includes tumor cells[14]. Pembrolizumab is a selective humanized monoclonal antibody of the immunoglobulin G4/κ type, designed to specifically hinder the interaction between PD-1 and PD-L1/PD-L2 by directly binding to PD-1[15]. Previous studies have shown that checkpoint inhibitors targeting PD-1 and cytotoxic T-lymphocyte-associated protein 4 (CTLA-4), including nivolumab and ipilimumab, respectively, synergize and stimulate an immune response against tumors through separate but complementary mechanisms. Nivolumab, a human monoclonal antibody blocking PD-1, has received accelerated approval from the U.S. Food and Drug Administration (FDA) for use either alone or in combination with low-dose ipilimumab in patients diagnosed with MSI-H or dMMR mCRC. Another findings also reported that both pembrolizumab and nivolumab, inhibitors targeting PD-1, have demonstrated significant antitumor efficacy in patients with previously treated MSI-H/dMMR metastatic CRC[7–9].

Due to their distinct mechanism of action, immune checkpoint inhibitors are linked to adverse events (AEs) that contrast with those typically associated with chemotherapy. These immune-related AEs commonly impact various bodily systems including the skin, gastrointestinal (GI) tract, lungs, kidneys, endocrine glands, and liver. Timely detection and appropriate management of these AEs, which may involve the use of systemic corticosteroids as necessary, could enhance outcomes for patients undergoing checkpoint inhibitor therapy (Brahmer et al., 2018). Understanding the predictors of response and resistance to immunotherapy is crucial for identifying patients who are most likely to benefit from these agents. Pembrolizumab, Nivolumab, and Ipilimumab represent novel therapeutic options for patients with advanced colorectal cancer, offering the potential for durable responses and improved survival outcomes. Therefore, this systematic review and meta-analysis aims to provide a comprehensive overview of the efficacy and safety profiles of Pembrolizumab, Nivolumab, and Ipilimumab in advanced colorectal cancer, with insights into future directions and challenges in the immunotherapy landscape.

## Material and Method

### Registration

The systematic review and meta-analysis titled “The Efficacy and Safety of Pembrolizumab, Ipilimumab, and Nivolumab Monoteraphy and Combination for Advanced Colorectal Cancer: A Systematic Review and Meta-Analysis” was officially registered on the Prospero on 24 February 2024 with registration number: CRD42024512511

### Eligibility criteria

This review adhered to the guidelines stipulated by the Preferred Reporting Items for Systematic Reviews and Meta-Analyses (PRISMA) statement and the Cochrane Handbook[16,17]. The inclusion criteria encompassed studies involving patients of any age diagnosed with advanced or metastatic colorectal cancer, regardless of microsatellite instability-high and/or mismatch repair-deficient (MSI-H/ dMMR) status, treated by Pembrolizumab, Ipilimumab, and Nivolumab as the monotherapy or combination therapy. The primary focus was assessing the efficacy and safety of this treatment. The criteria for inclusion necessitated the availability of data on the mentioned primary outcomes published from 2014 until 2024. Furthermore, the eligible studies were restricted to observational designs (cohort/case-control) or randomized clinical trials (RCTs) or clinical trial.

The exclusion criteria were technical reports, editor responses, narrative reviews, systematic reviews, meta-analyses, non-comparative research, in silico studies, in vitro studies, in vivo studies, scientific posters, study protocoals, and conference abstracts. For data synthesis, the study will be grouped based on relevant comparators that include the use of Nivolumab+Ipilimumab, Nivolumab, and Pembrolizumab monotherapy.

### Literature Search and Study Selection

A comprehensive search of English literature across eight international databases— Scopus, PubMed, Cochrane Library, Sage pub, ProQuest, Science Direct, Research Gate, Epistemonikos—was conducted by a team of 12 independent authors from inception until February 1st, 2024. To capture all potentially relevant literature, a combination of keywords was utilized, employed the following keywords: “Pembrolizumab” or “Nivolumab” or “Ipilimumab” and “colorectal carcinoma” and “therapeutic”. The studies were organized and managed using the Mendeley Group Reference Manager in the authors’ library. Initially, duplicate articles were removed, followed by screening based on titles and abstracts. Each researchers independently reviewed the titles and abstracts, resolving any disparities through discussion until reaching an agreement. Subsequently, they collaborated in 6 pairs of group to review the titles and abstracts of all retrieved articles. If necessary, the corresponding author was involved to make the final decision. Articles that passed this initial screening underwent full-text evaluation against predetermined inclusion/exclusion criteria. The full-text screening was conducted by the same 12 independent researchers with any discrepancies resolved through discussion with the corresponding author as the final decision maker.

**Table 1.**
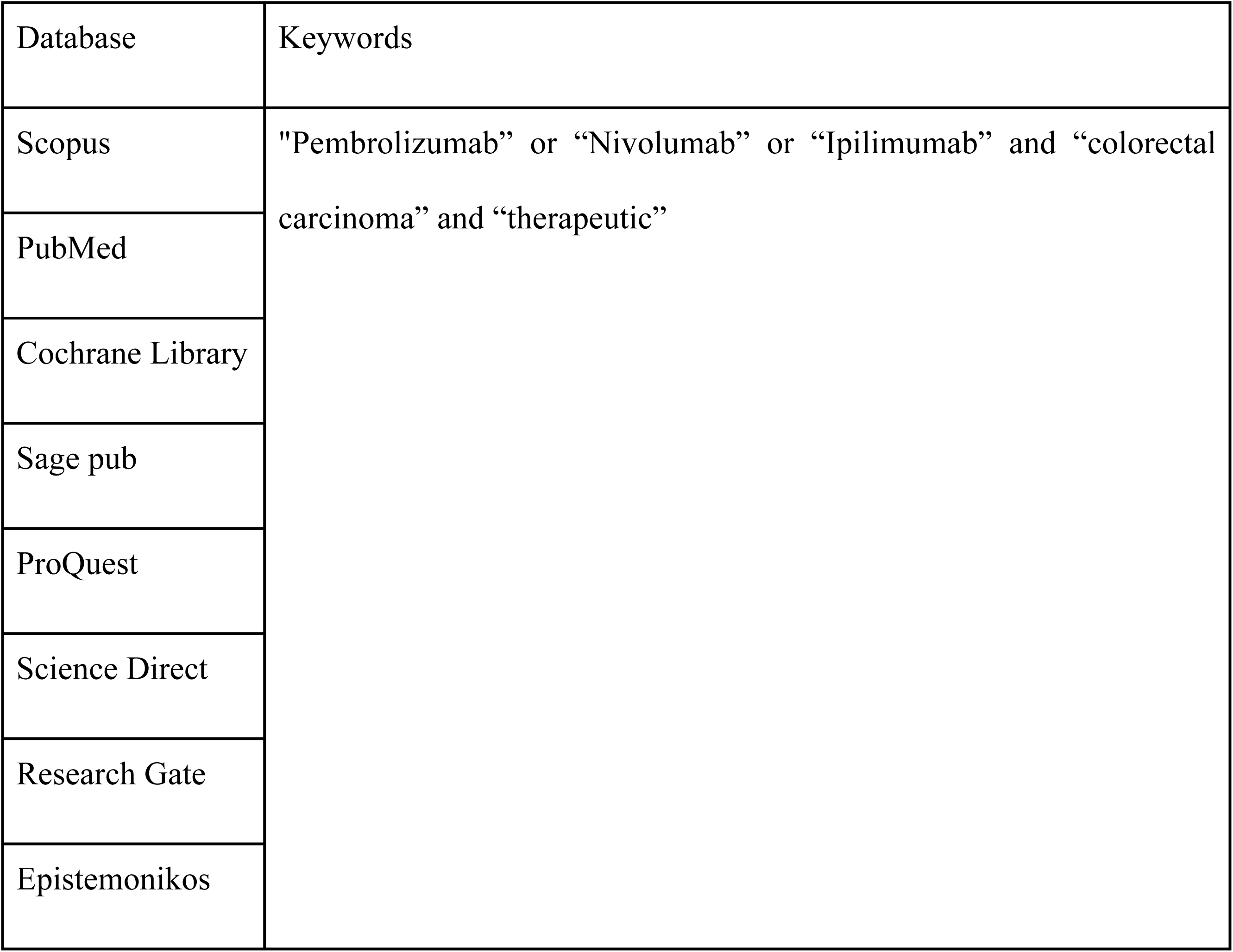
Keyword Used in Literature Searching.

### Data Extraction

The following data were extracted for analytical purposes: study ID, publication year, country, study design, sample size, baseline characteristics of participants (mean age and sex distribution), follow-up duration, the number of participants in the buried and exposed K-wire groups, and the outcomes of interest. These data, extracted by 12 independent authors, were tabulated using Microsoft Excel 2019. The primary outcome assessment of efficacy incorporated into two general parameters, survival and therapy response. The survival parameter is indicated as overall survival (OS), progression-free survival (PFS) (The duration spanning both the treatment period and the subsequent phase following the intervention for a medical condition, such as cancer, wherein a patient sustains a state in which the disease neither progresses nor deteriorates). Furthermore, the response evaluation was assessed by objective response rate (ORR) These parameters were chosen to holistically evaluate the influence of the therapeutic intervention among individuals afflicted with colorectal cancer. The adverse effect occured during the treatment indicated as our secondary outcomes.

### Risk of Bias Assessment

The evaluation of bias risk was carried out by five independent authors utilizing validated instruments. To assess the quality of the included Randomized Controlled Trials (RCTs), the Cochrane Collaboration’s Risk of Bias version 2 (RoB v2) tool was employed, comprising methodological assessments across five domains: (a) randomization process; (b) deviations from intended interventions; (c) missing outcome data; (d) measurement of the outcome; and (e) selection of the reported results. The outcomes are depicted in Fig 2. The authors’ appraisals were categorized as “low risk,” “high risk,” or “some concerns” of bias. Two review authors (KCT, DR) independently utilized the tool for each included study and documented supporting information for their assessments of risk of bias in each domain. In evaluating the quality of included cohort/case-control studies or non randomized clinical trial, the Risk Of Bias In Non-randomised Studies - of Interventions (ROBINS-I) was utilized, covering seven domains: (1) confounding; (2) selection of participants; (3) classification of intervention; (4)deviation from intended intervention; (5) missing data; (6) measurement of outcomes; (7) selection of the reported result. The authors’ appraisals were categorized as “low risk,” “high risk,” or “some concerns” of bias. Two review authors (KCT, DR) independently utilized the tool for each included study and documented supporting information for their assessments of risk of bias in each domain. Any disparities in risk of bias assessments or justifications were addressed through discussion to achieve consensus between the two review authors ( KCT, DR) If needed, corresponding author (EM) served as an arbiter to facilitate resolution

### Statistical Analysis

For the analytical synthesis of continuous variable outcomes, we employed the mean difference (SMD) with 95% confidence intervals (95% CI), utilizing the Inverse-Variance formula. Dichotomous variable outcomes were pooled into risk ratios (RR) with 95% CI using the Mantel-Haenszel formula. Given the anticipated substantial heterogeneity due to variations in population characteristics and follow-up durations, random-effect models were selected for this review. Heterogeneity between studies was assessed using the I-squared (I2) statistic, where values exceeding 50% were categorized as significant heterogeneity. To facilitate pooled analysis, we employed a combined formula from Luo D et al.[15] and Wan X et al.[16] for transforming data expressed as the median and interquartile range (IQR), or median, minimum, and maximum into mean and standard deviations (SD).

### Reporting bias and Certainity assessment

Publication bias analysis was conducted when there were more than 10 studies for each outcome of interest. In cases of funnel plot asymmetry, a thorough examination of characteristics aimed to discern whether observed asymmetry could be attributed to publication bias or other factors like methodological heterogeneity among studies. All statistical analyses were executed using JAMOVI 2.3. The robustness of outcomes was scrutinized through sensitivity meta-analysis, exclusively considering studies with an overall low risk of bias.

In the subgroup analysis, a commonly employed statistical methodology for data analysis involves elucidating associations between a dependent variable and one or more independent variables. Utilizing linear regression, the model centers on the dependent variable (e.g., sample size, age, sex, and follow-up duration), evaluating whether these variables influence the observed associations. The selection of variables is guided by theoretical considerations and their relevance to analytical objectives. JAMOVI 2.3 was employed for conducting subgroup analysis, ensuring robust data interpretation.

## Result

### Study Selection and Characteristics

A comprehensive literature search across eight international databases identified a total of 27,974 studies. Then the filter of released date and type of article were performed, 15,955 articles were automatically removed before being included into screening process. After the elimination of duplicate entries and initial screening based on titles and abstracts, 11,519 studies were removed due to irrelevant topics and 458 studies were removed due to non-RCT and non-clinical trial studies, resulting in a final selection of 42 studies. Subsequently, a thorough evaluation of these 42 studies in full-text format revealed that 24 studies did not meet the desired outcomes and 5 studies were not retrievable. Consequently, only 13 studies satisfied the eligibility criteria and were included in the final analysis, as illustrated in Figure 1. Sample sizes ranged from 11 to 307, with follow-up durations varying from 5.3 month to 44.5 month. A comprehensive overview of the baseline characteristics of the included studies is presented in Table 2.

**Figure 1.**
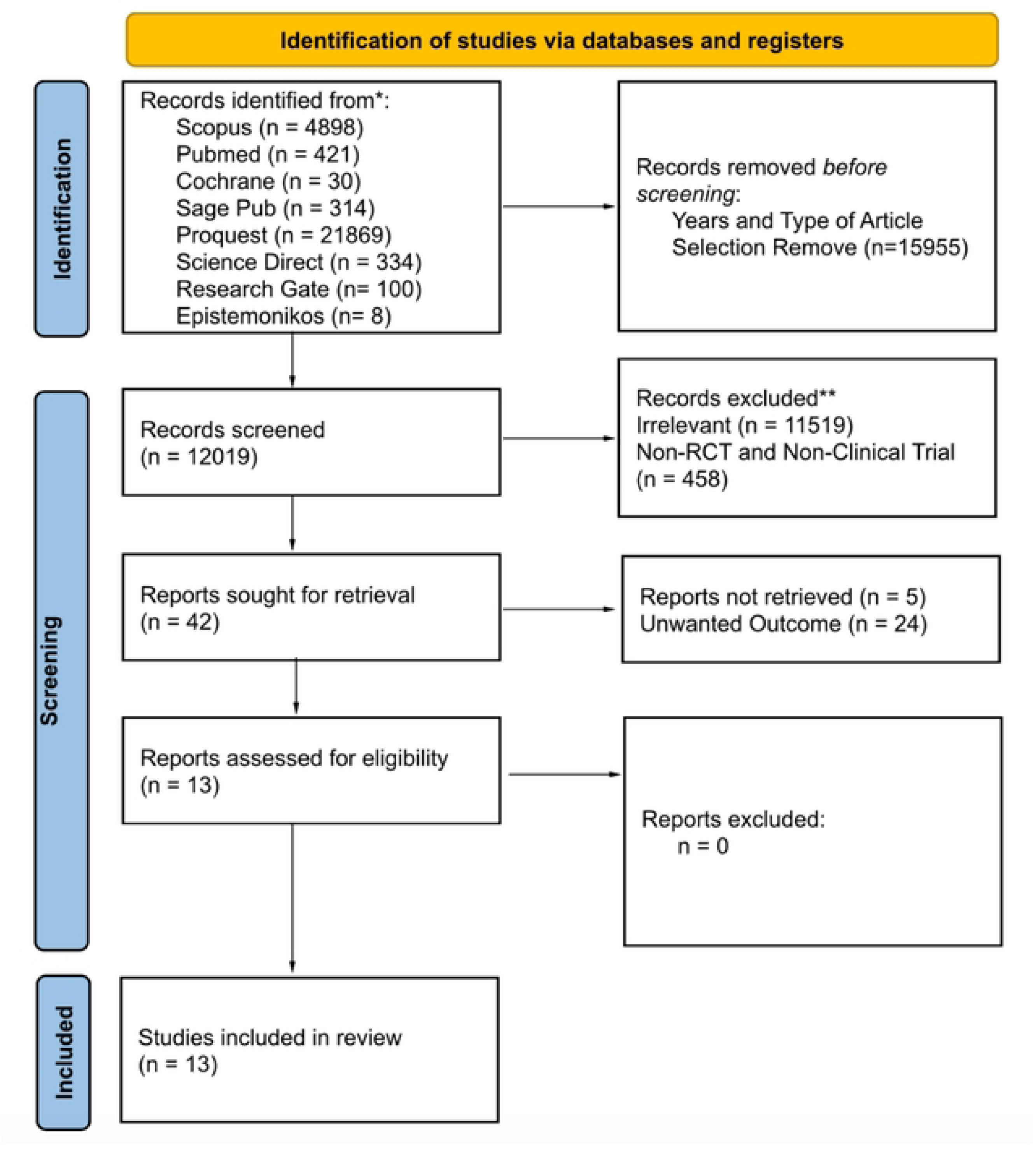
PRISMA 2020 flow diagram.

**Table 2.**
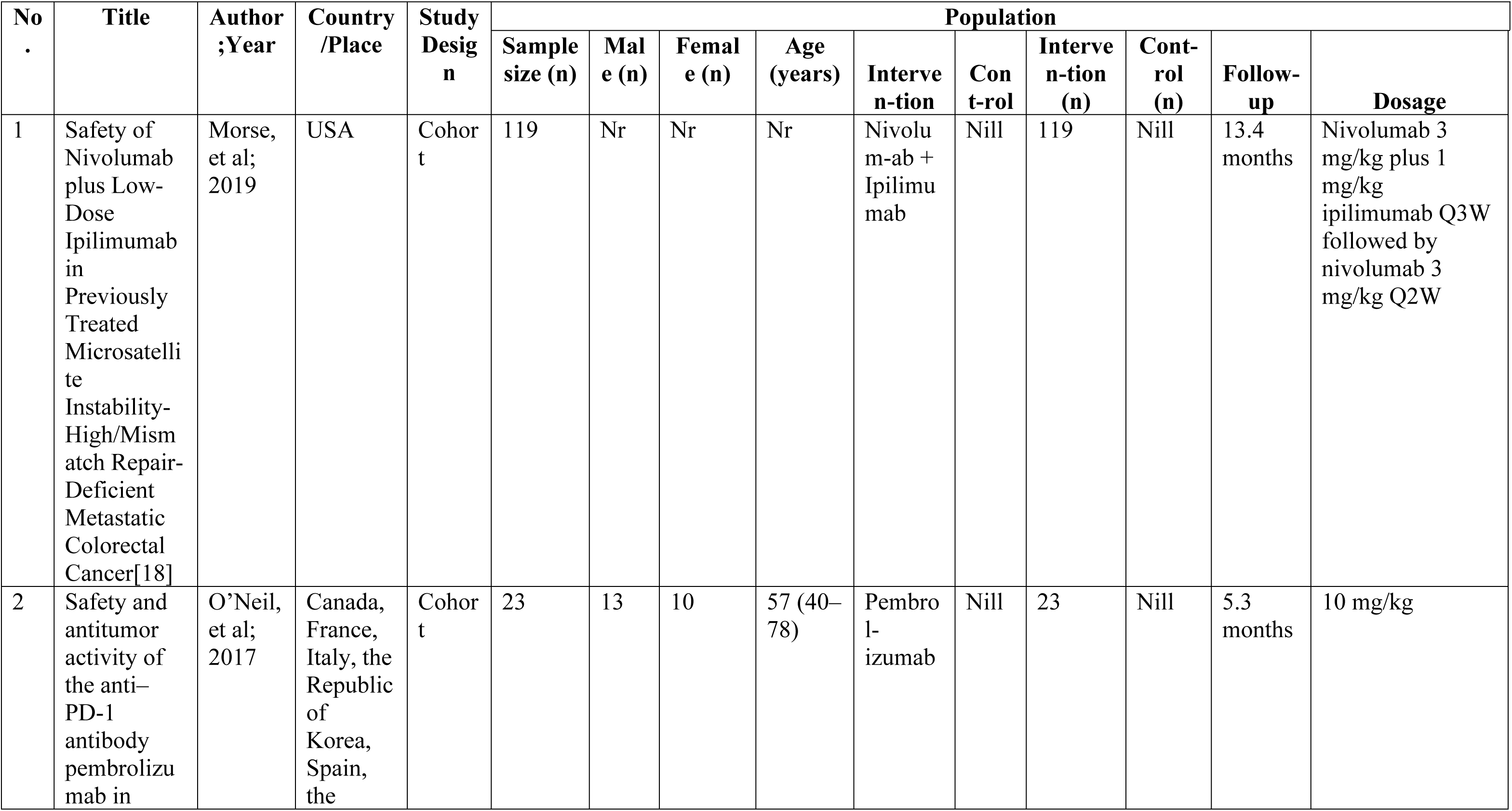
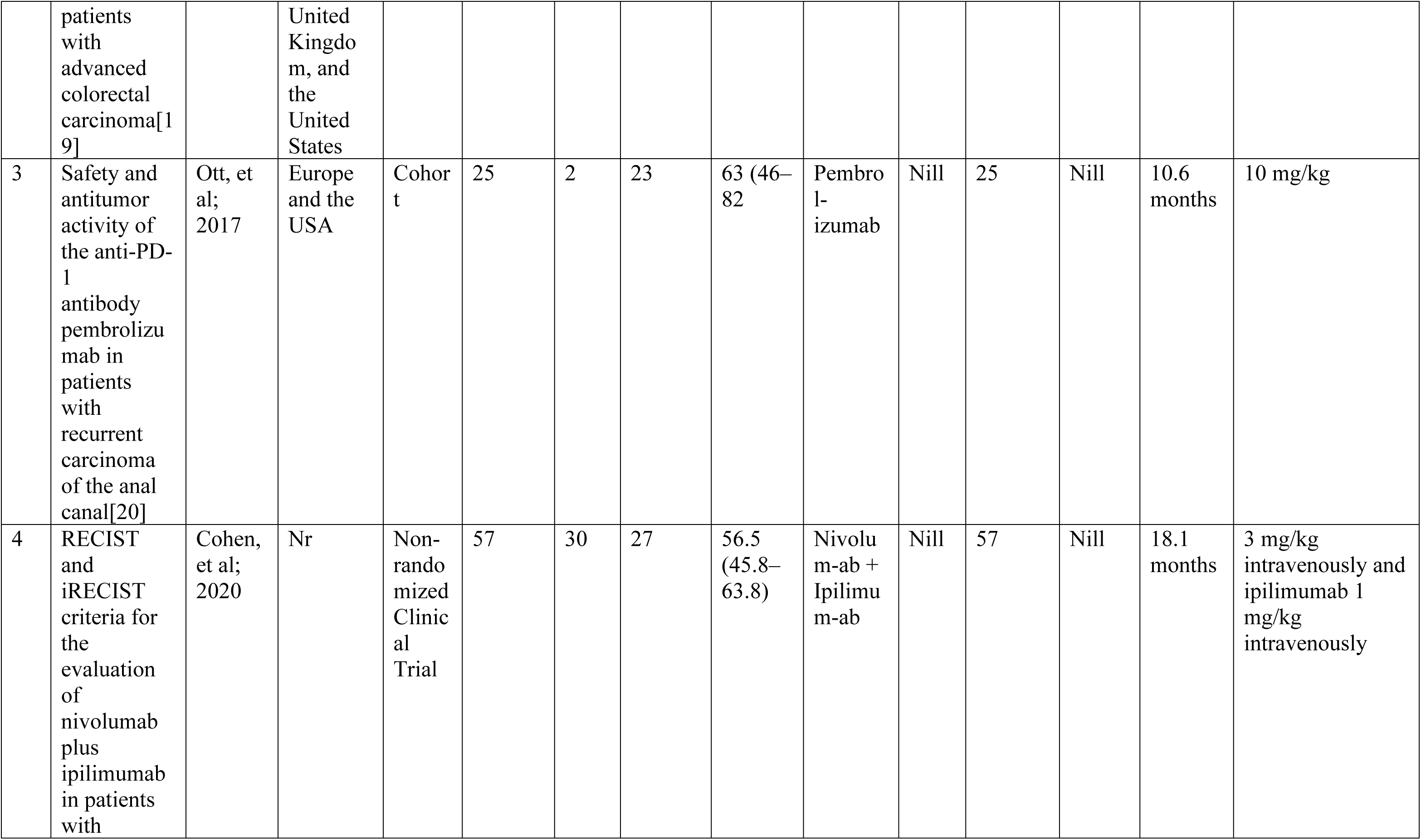
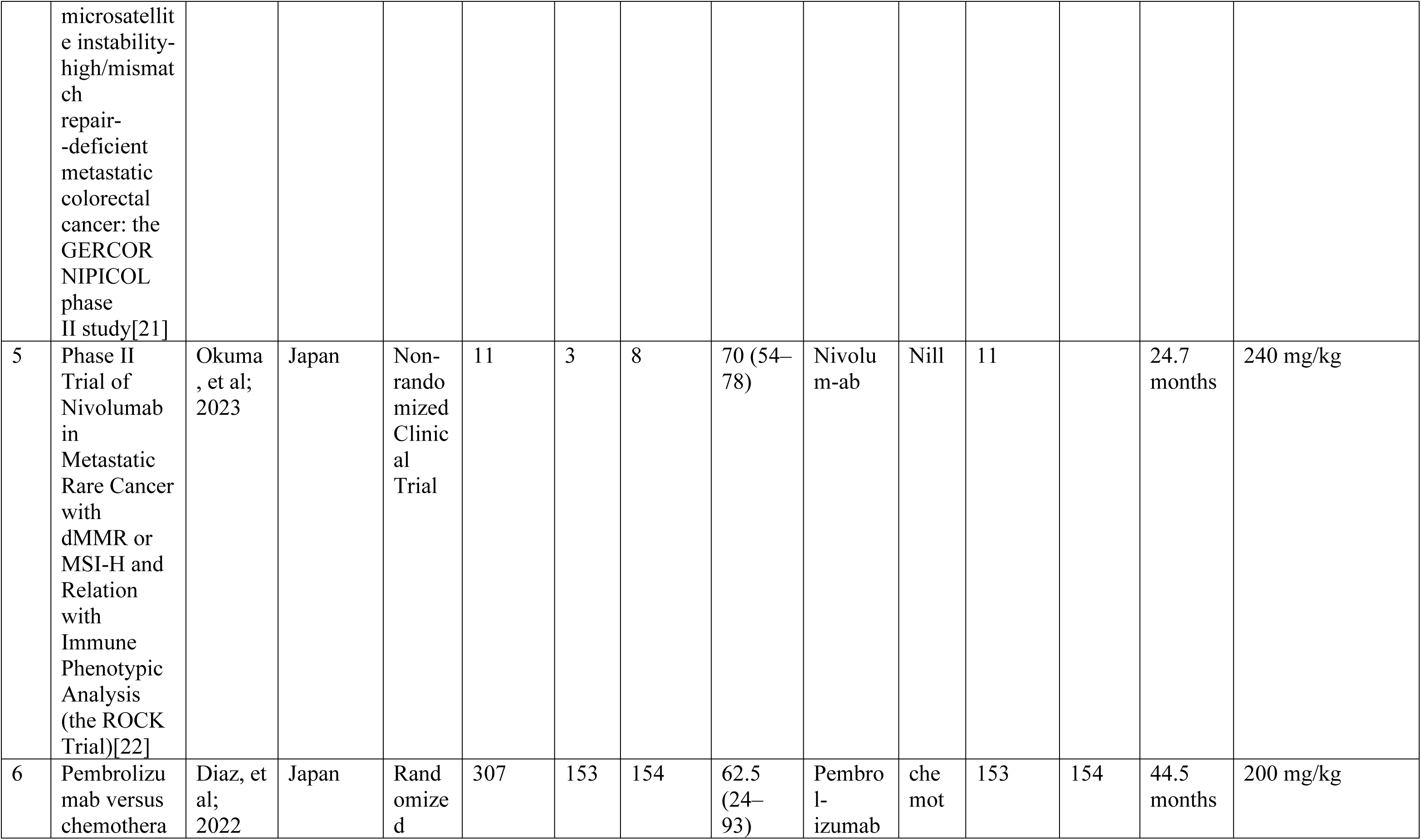
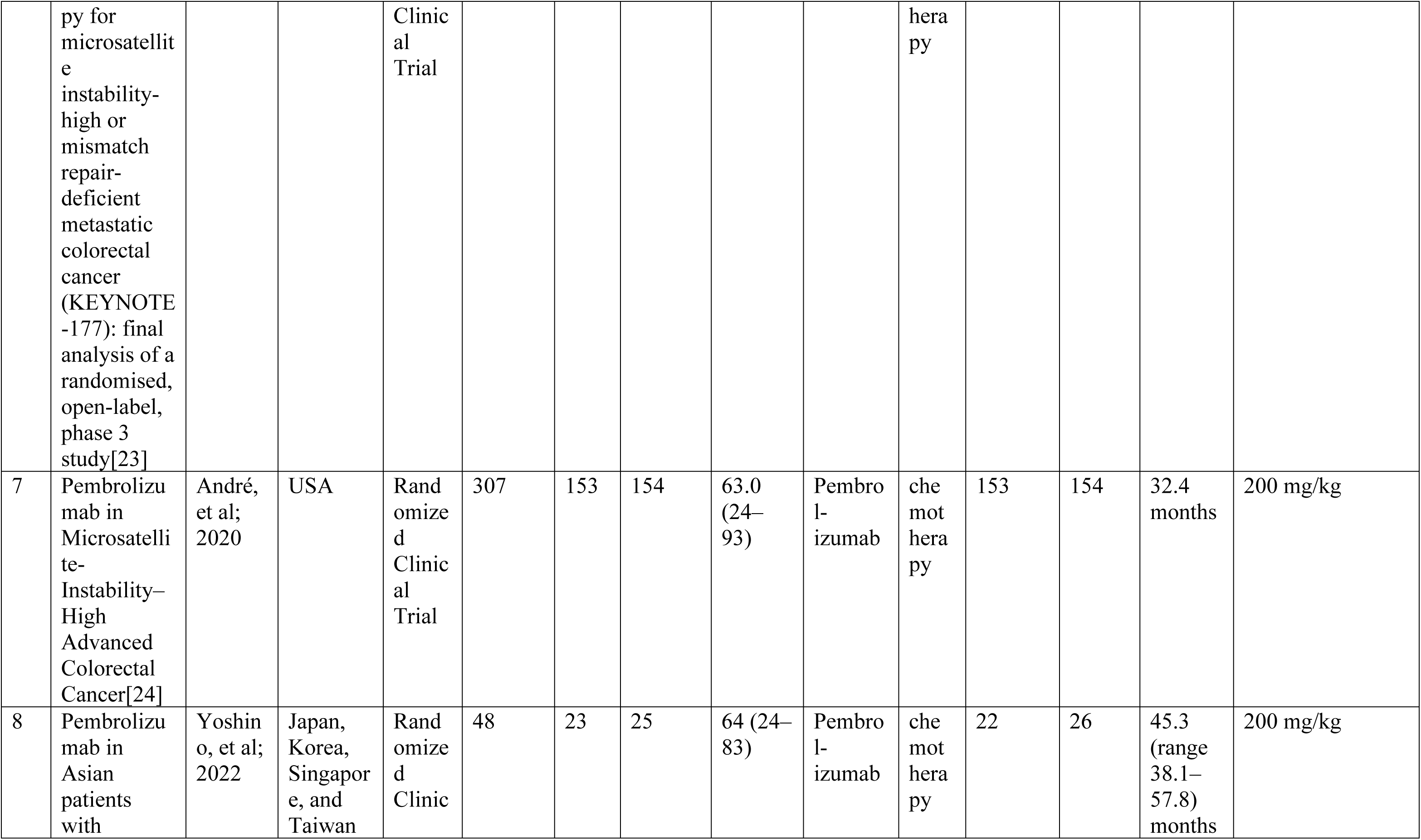
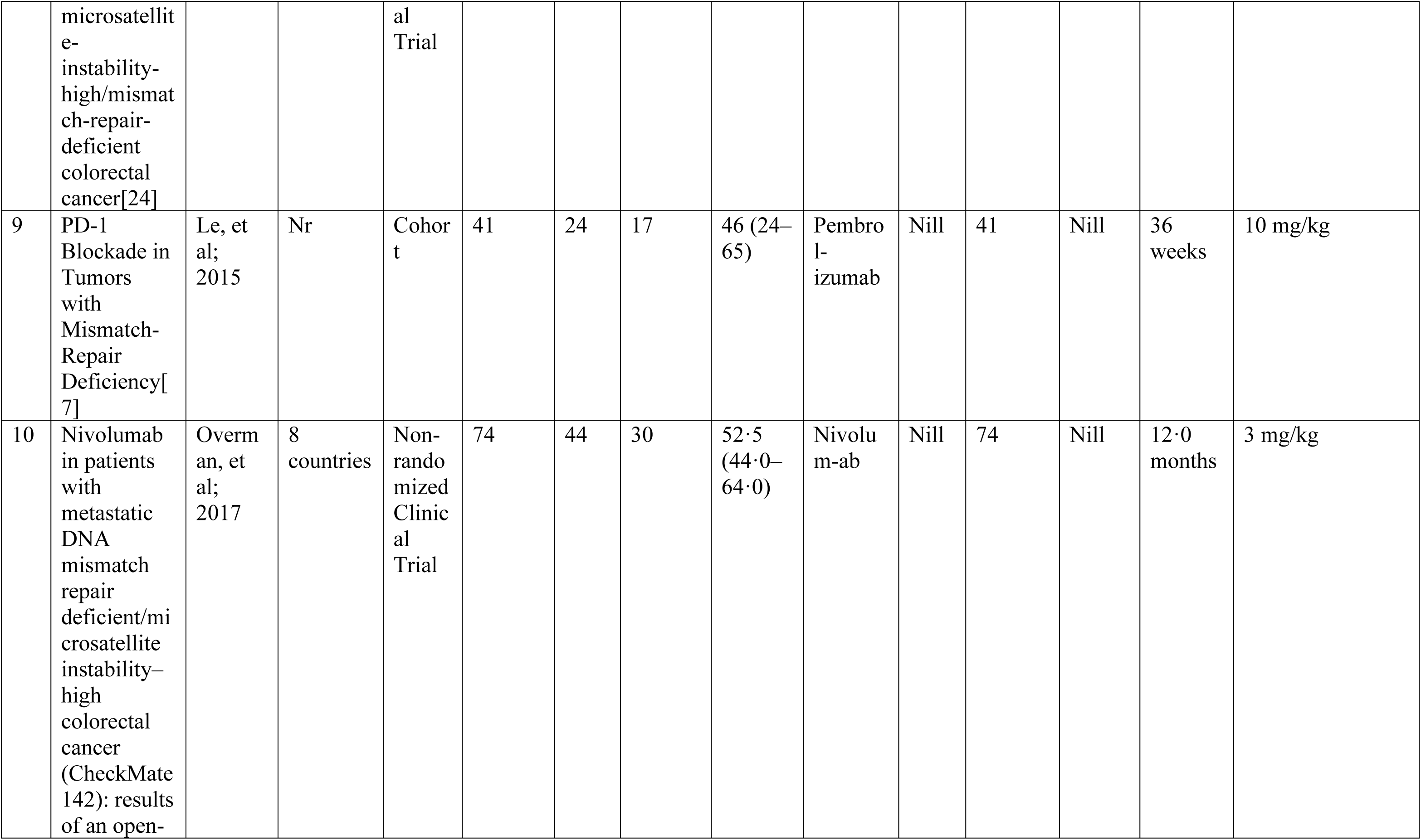
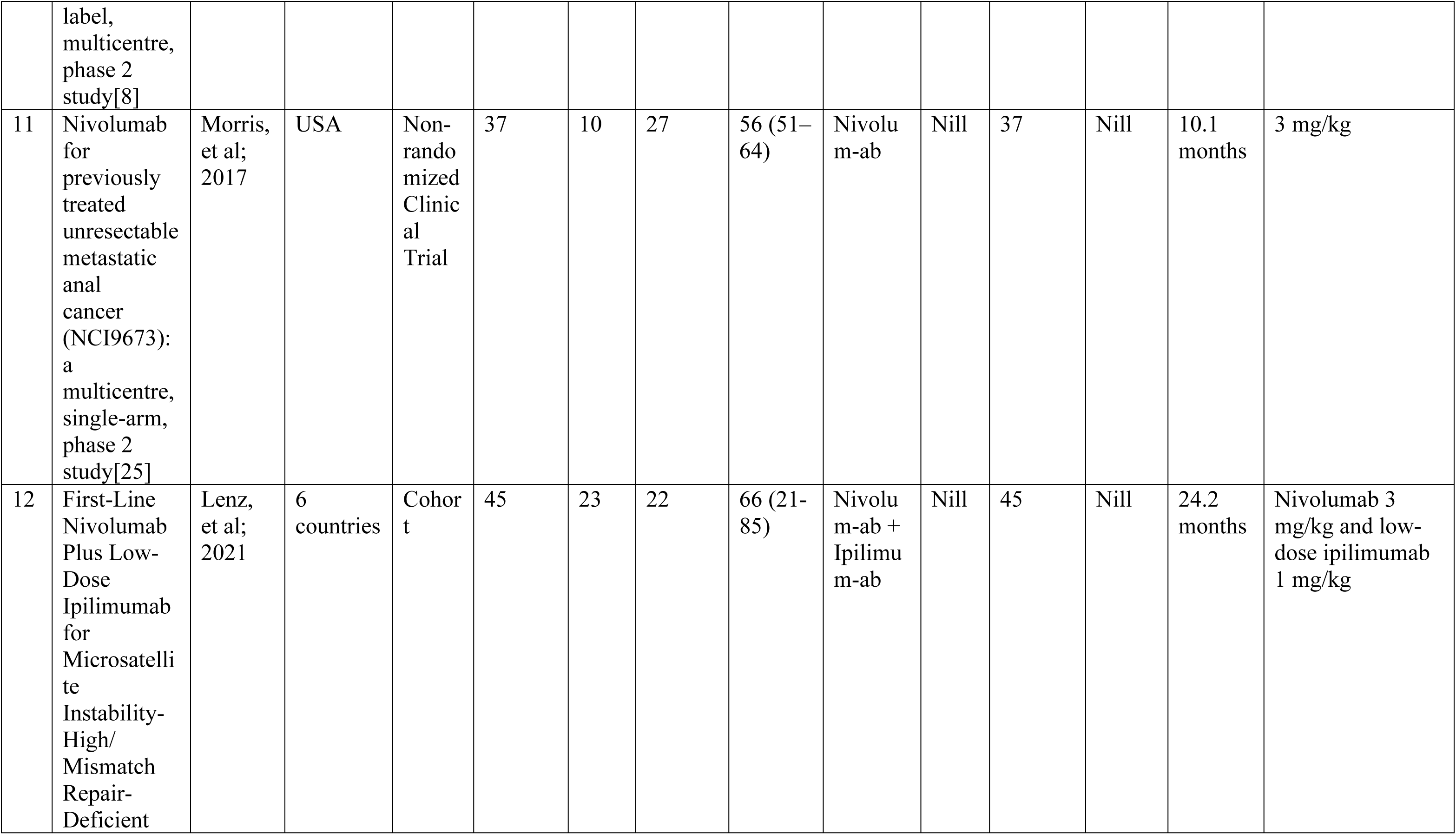
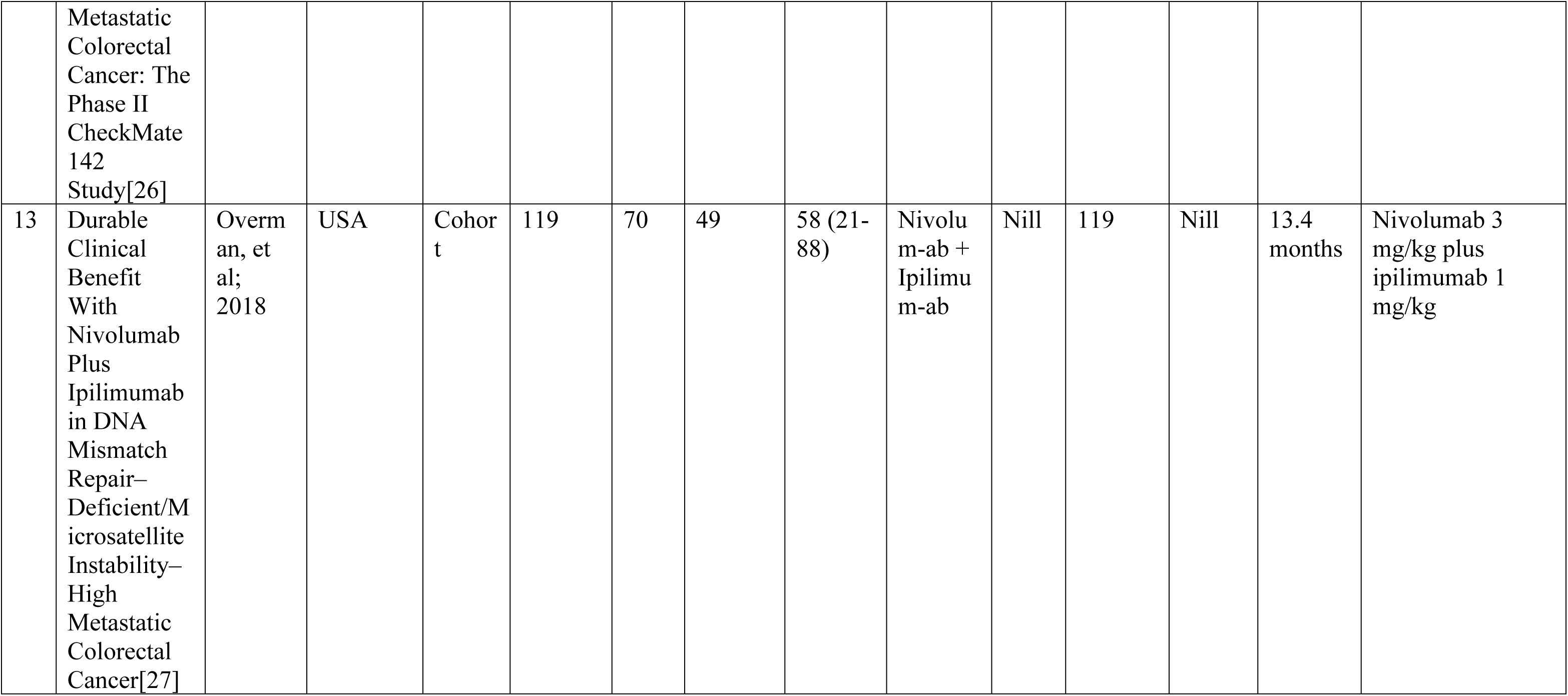
Study Characteristic.

### Risk of Bias in Studies

Each clinical trial and randomized controlled trial underwent a rigorous evaluation employing the ROB-2 method. Among the identified studies, three exhibited a level of bias concern, primarily attributed to confounding, classification of interventions, missing data, and reported results. The visualization of the risk-of-bias assessment is presented in Figure 2.

**Figure 2.**
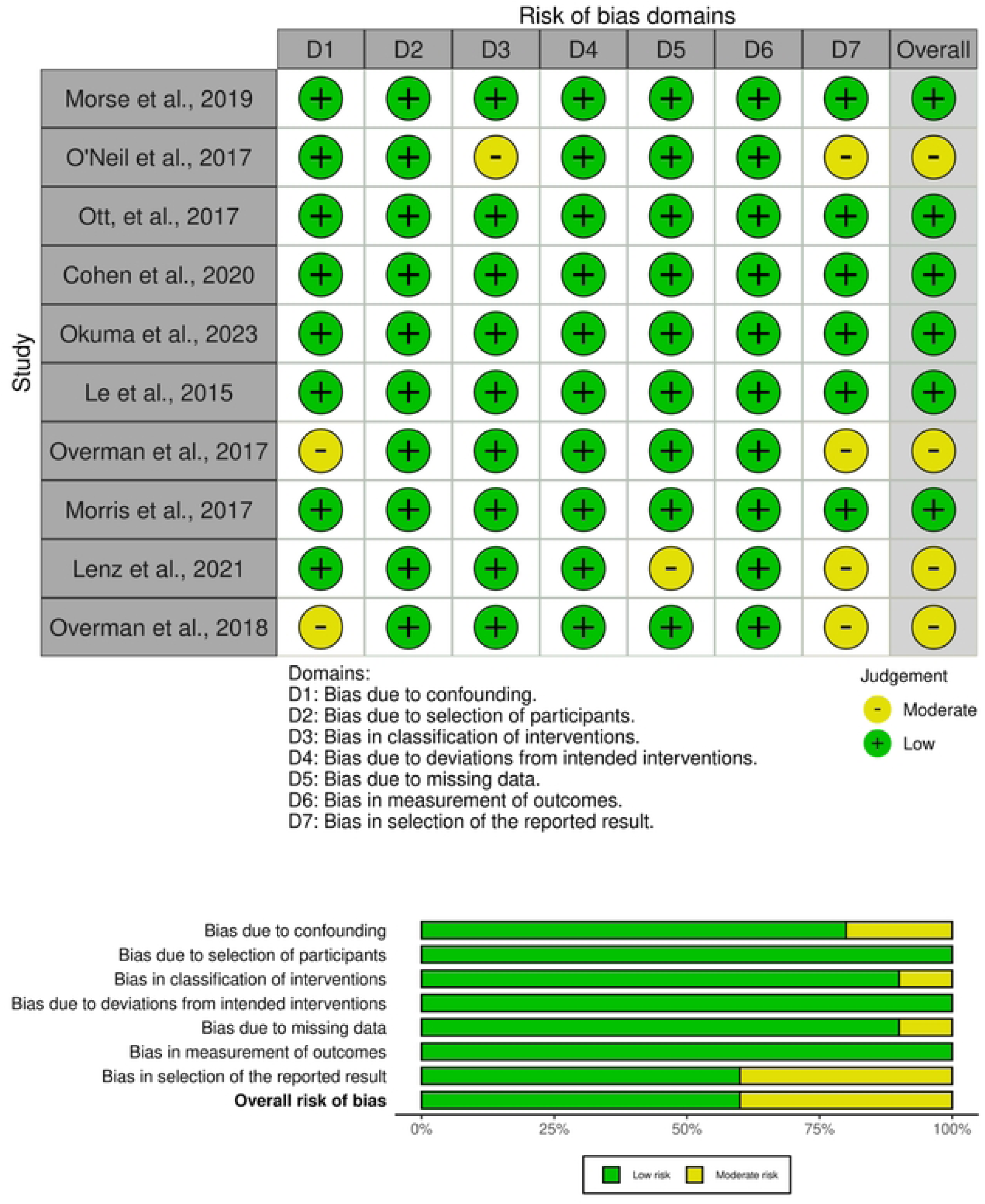
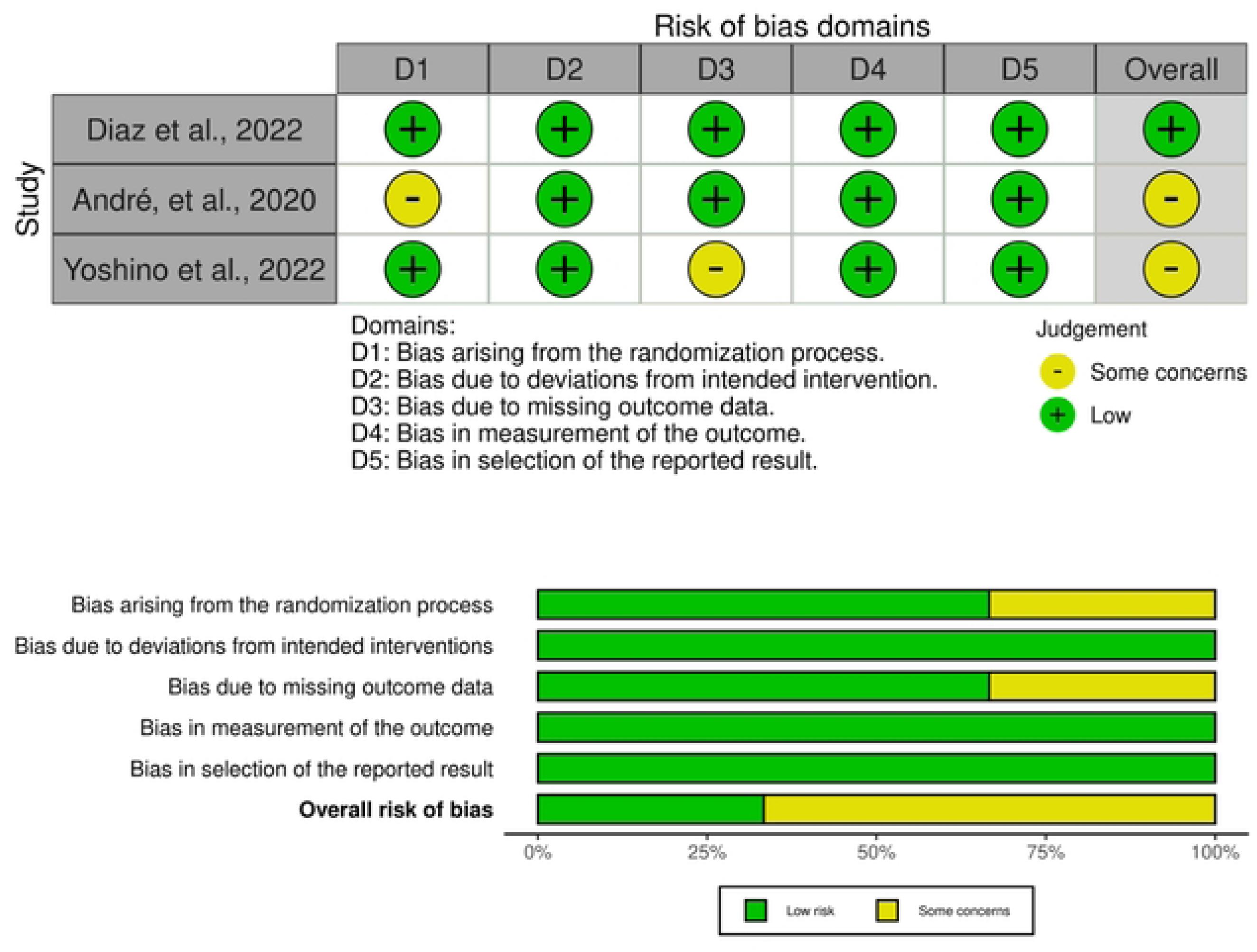
(A) ROBINS-I Risk of Bias Assessment; (B) ROB-2 Risk of Bias Assessment.

### Objective response rate (ORR)

Three studies that administered Nivolumab reported ORR with an effect size of **0.34 [95% CI 0.18; -0.49, P<0.001]**. Heterogeneity was found to be moderate and the funnel plot shows no evidence of heterogeneity.

**Figure 3.**
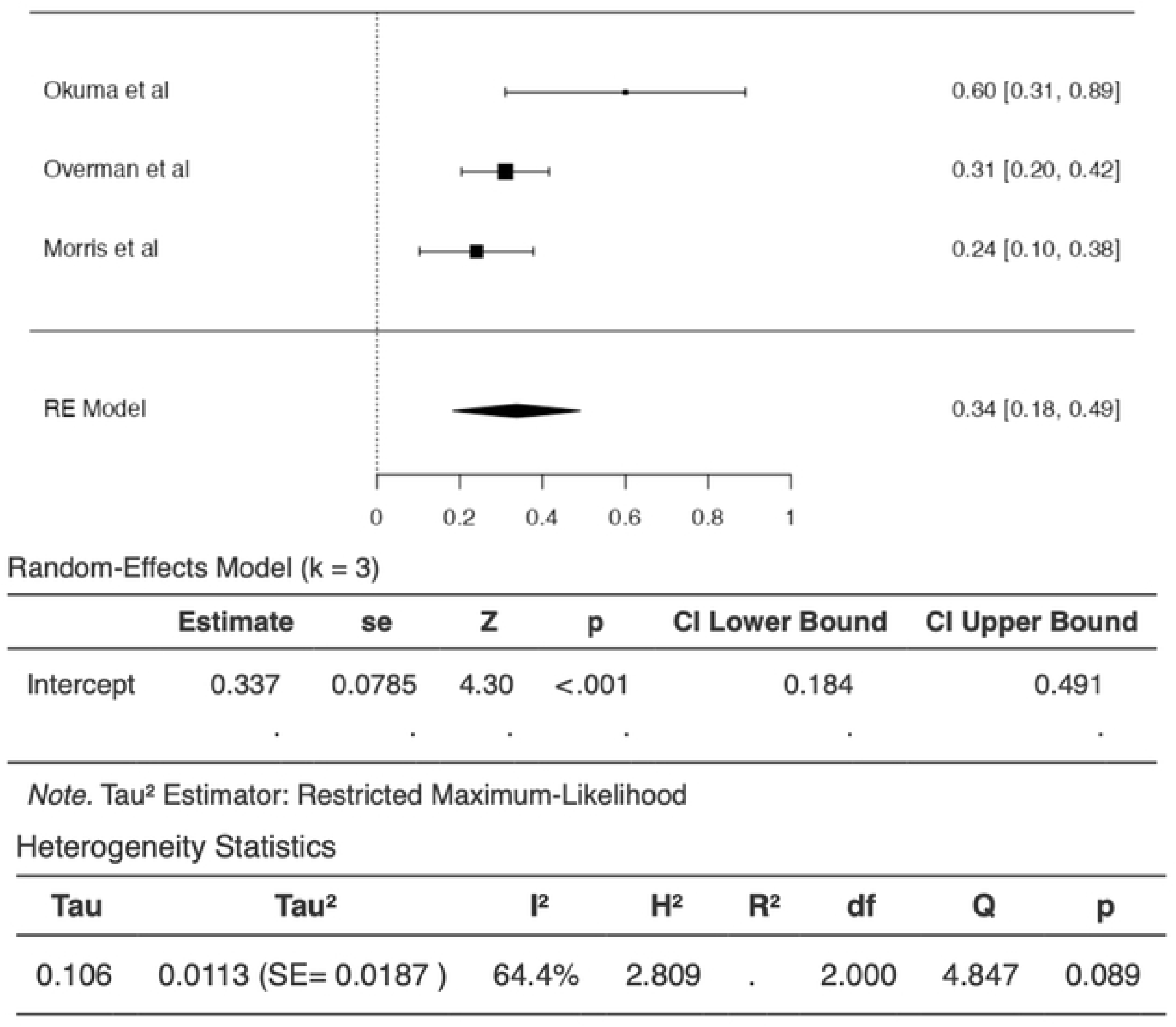
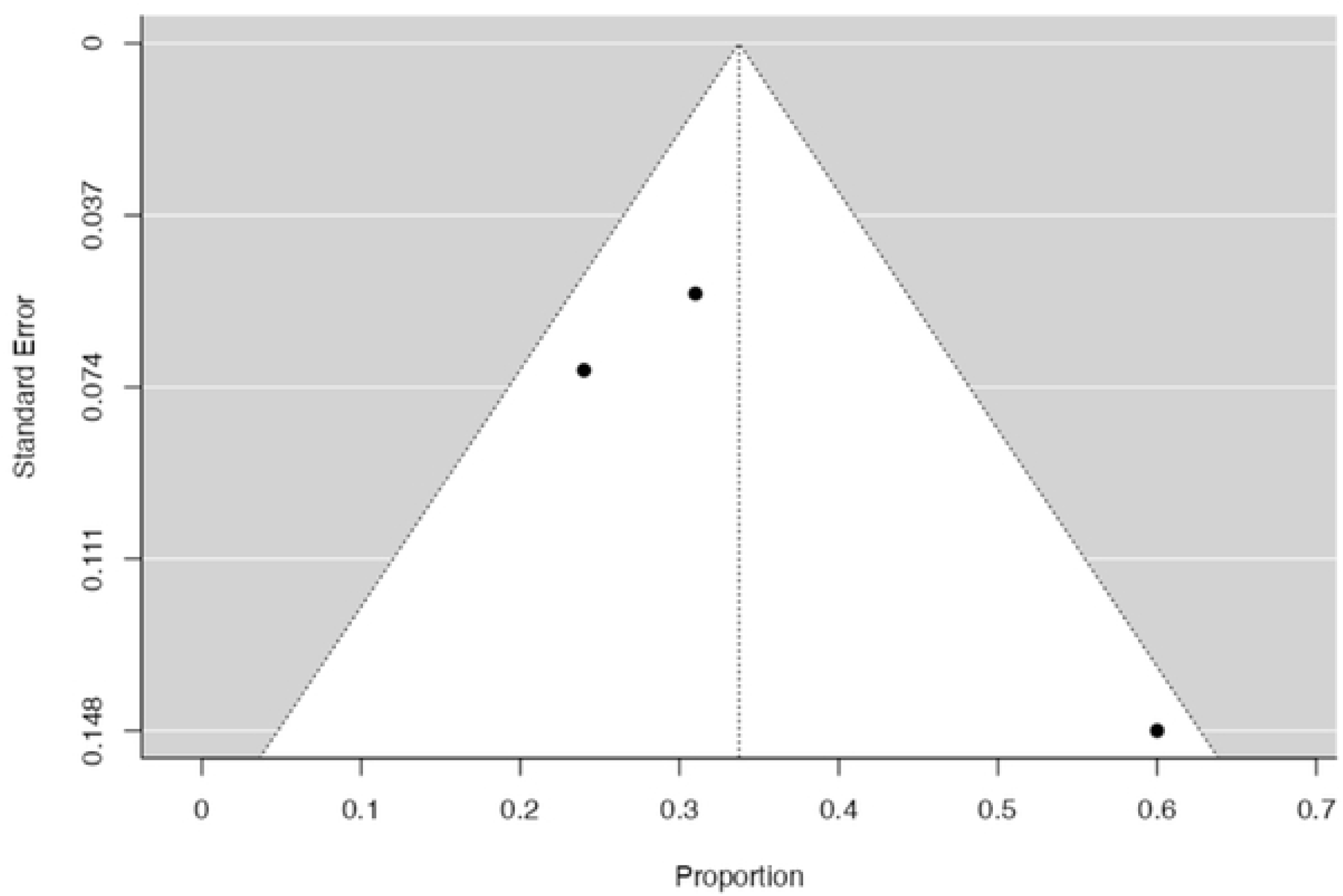
(A) Forest plot of ORR nivolumab intervention; (B) Funnel plot of ORR nivolumab intervention.

Four studies that administered Nivolumab and Ipilimumab reported ORR with an effect size of **0.53 [95% CI 0.42; -0.65, P<0.001]**. Heterogeneity was found to be high and the funnel plot shows evidence true of heterogeneity.

**Figure 4.**
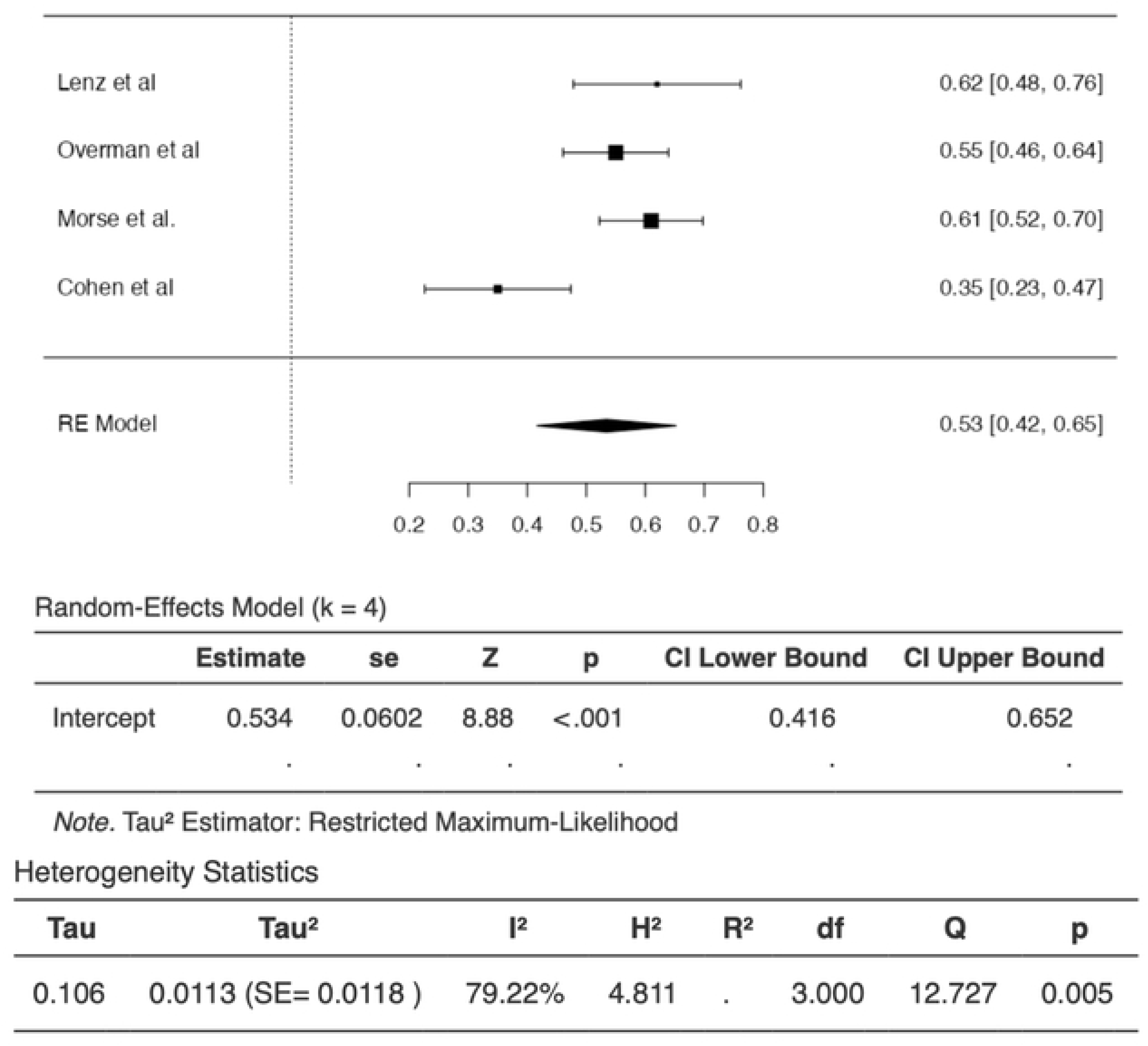
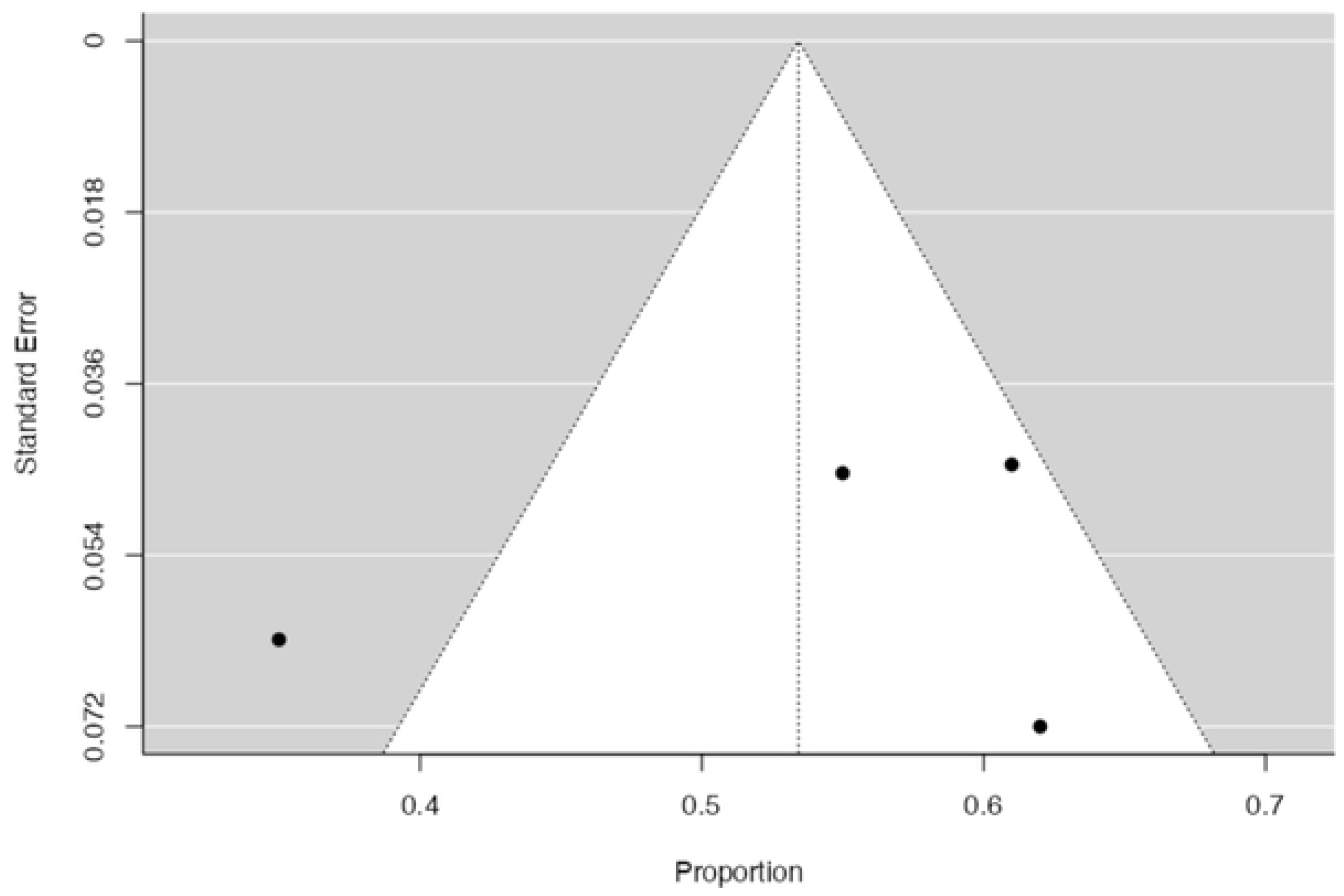
(A) Forest plot of ORR Nivolumab and Ipilimumab intervention; (B) Funnel plot of ORR Nivolumab and Ipilimumab intervention.

Six studies that administered Pembrolizumab reported ORR with an effect size of **0.34 [95% CI 0.19; 0.48, P<0.001]**. Heterogeneity was found to be high and the funnel plot shows evidence true of heterogeneity.

**Figure 5.**
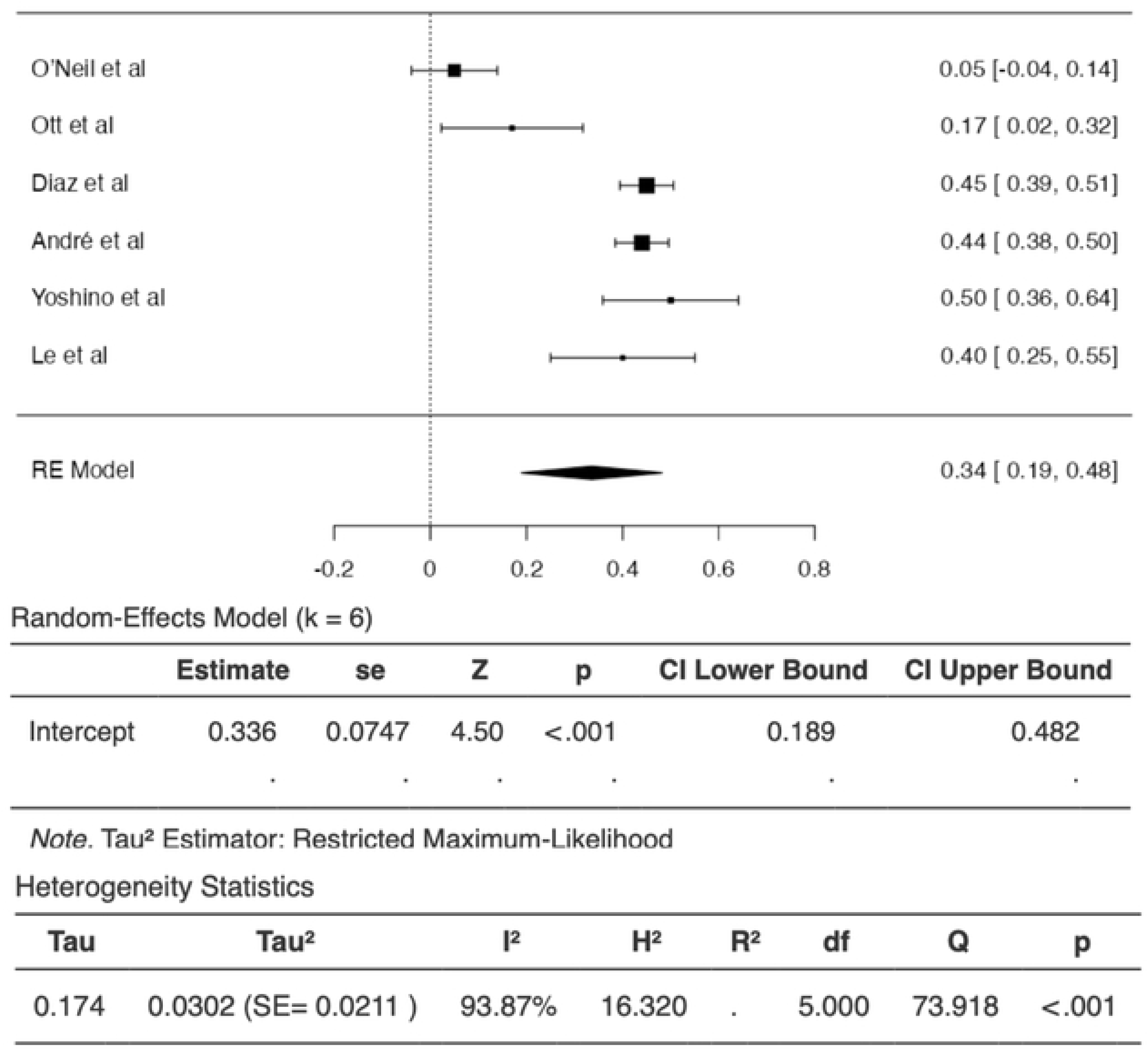
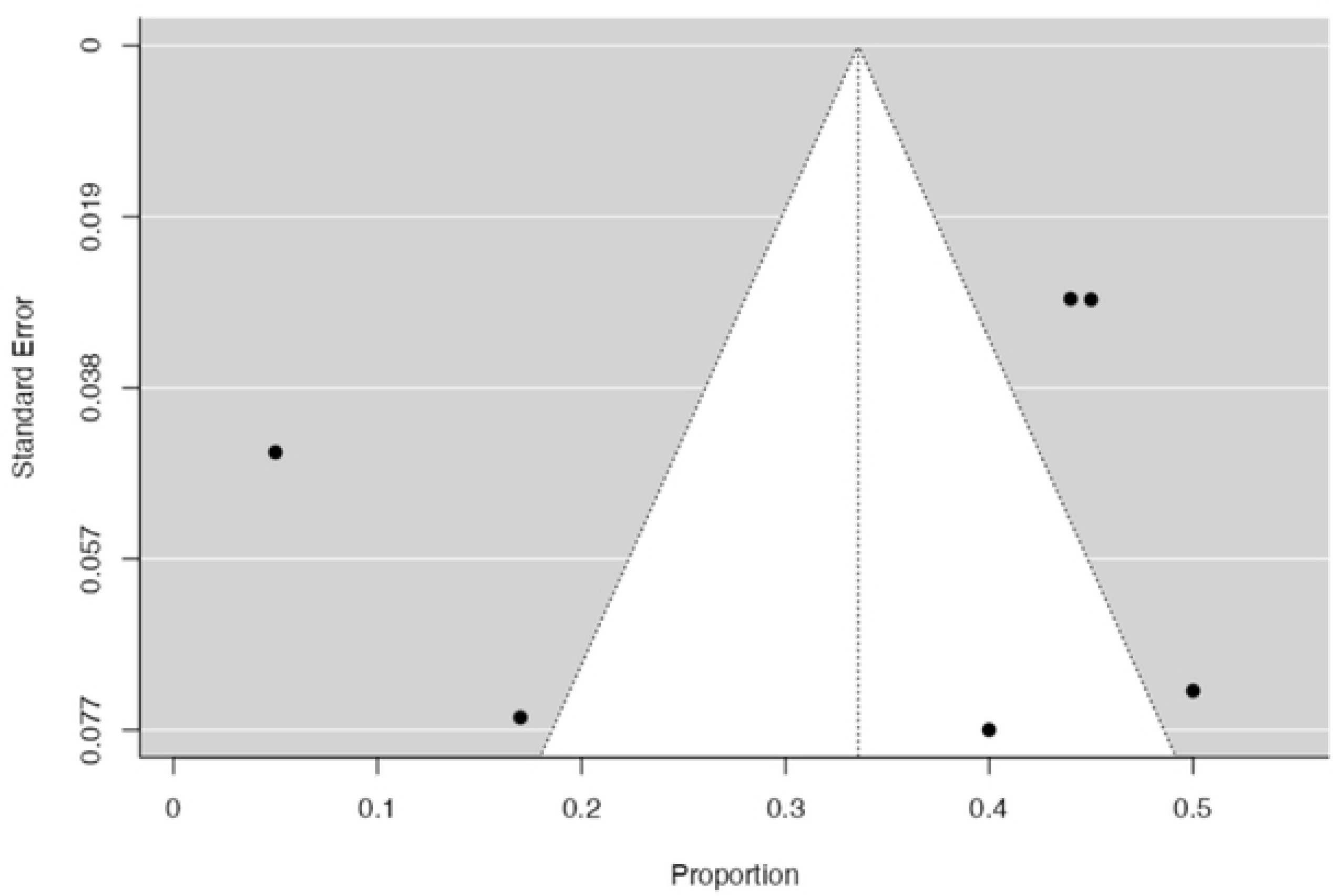
(A) Forest plot of ORR Pembrolizumab intervention; (B) Funnel plot of ORR Pembrolizumab intervention.

### Overall Survival

Three studies that administered Nivolumab reported OS with an effect size of **0.68 [95% CI 0.52; 0.84, P<0.001]**. Heterogeneity was found to be moderate and the funnel plot shows no evidence true of heterogeneity.

**Figure 6.**
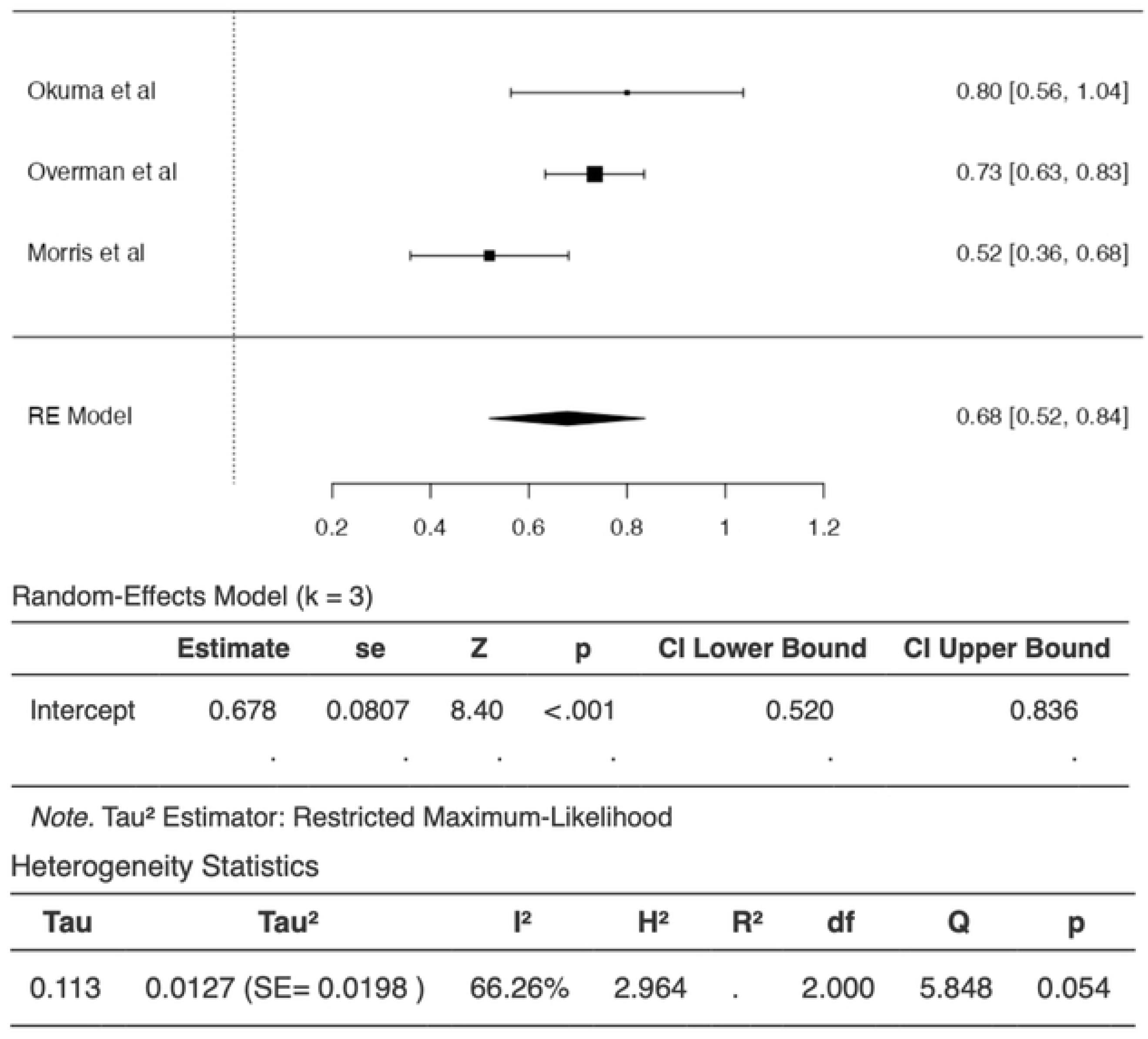
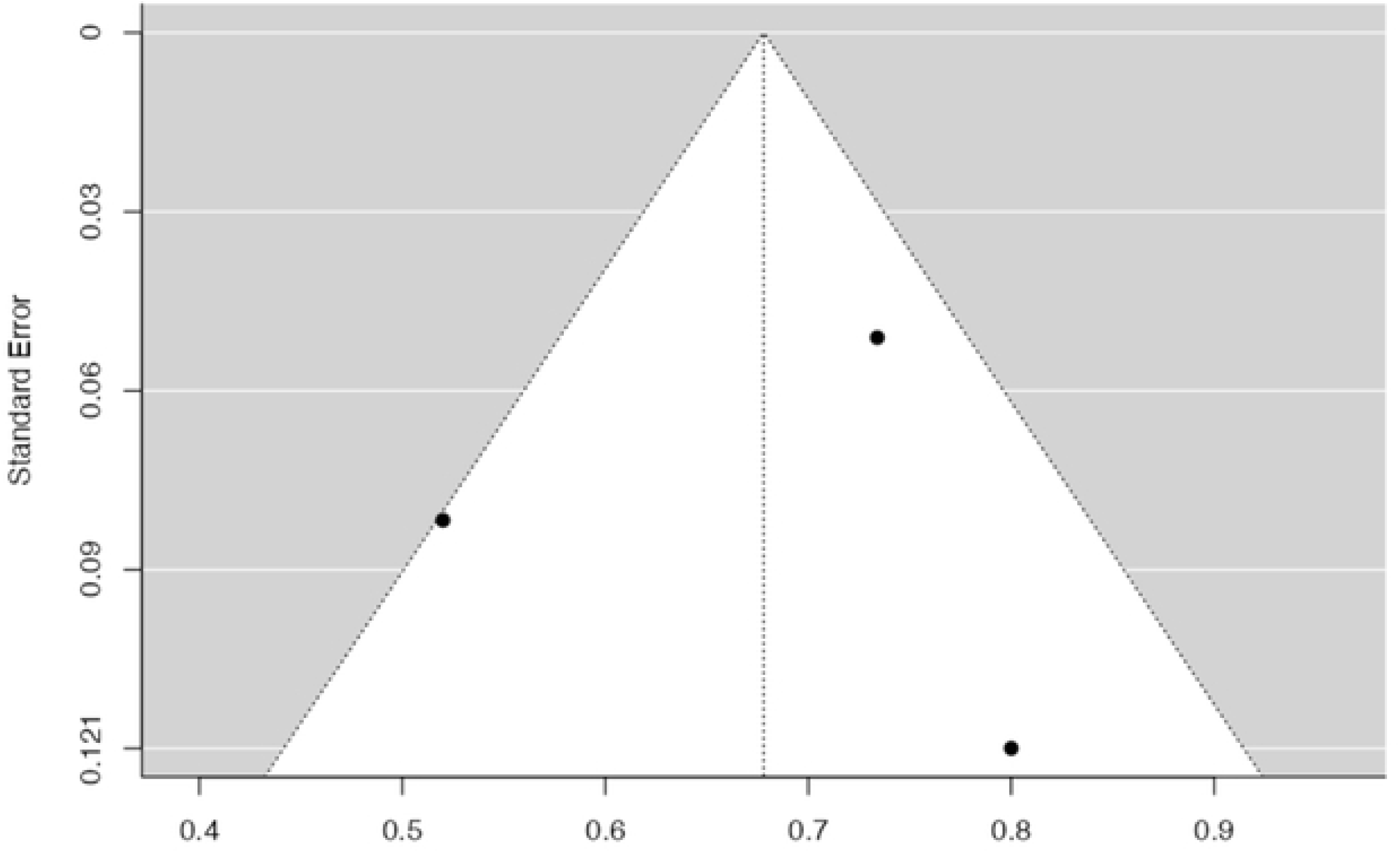
(A) Forest plot of OS Nivolumab intervention; (B) Funnel plot of OS Nivolumab intervention.

Four studies that administered Nivolumab and Ipilimumab reported OS with an effect size of **0.84 [95% CI 0.81; 0.88, P<0.001]**. Heterogeneity was found to be absent and the funnel plot shows no evidence true of heterogeneity.

**Figure 7.**
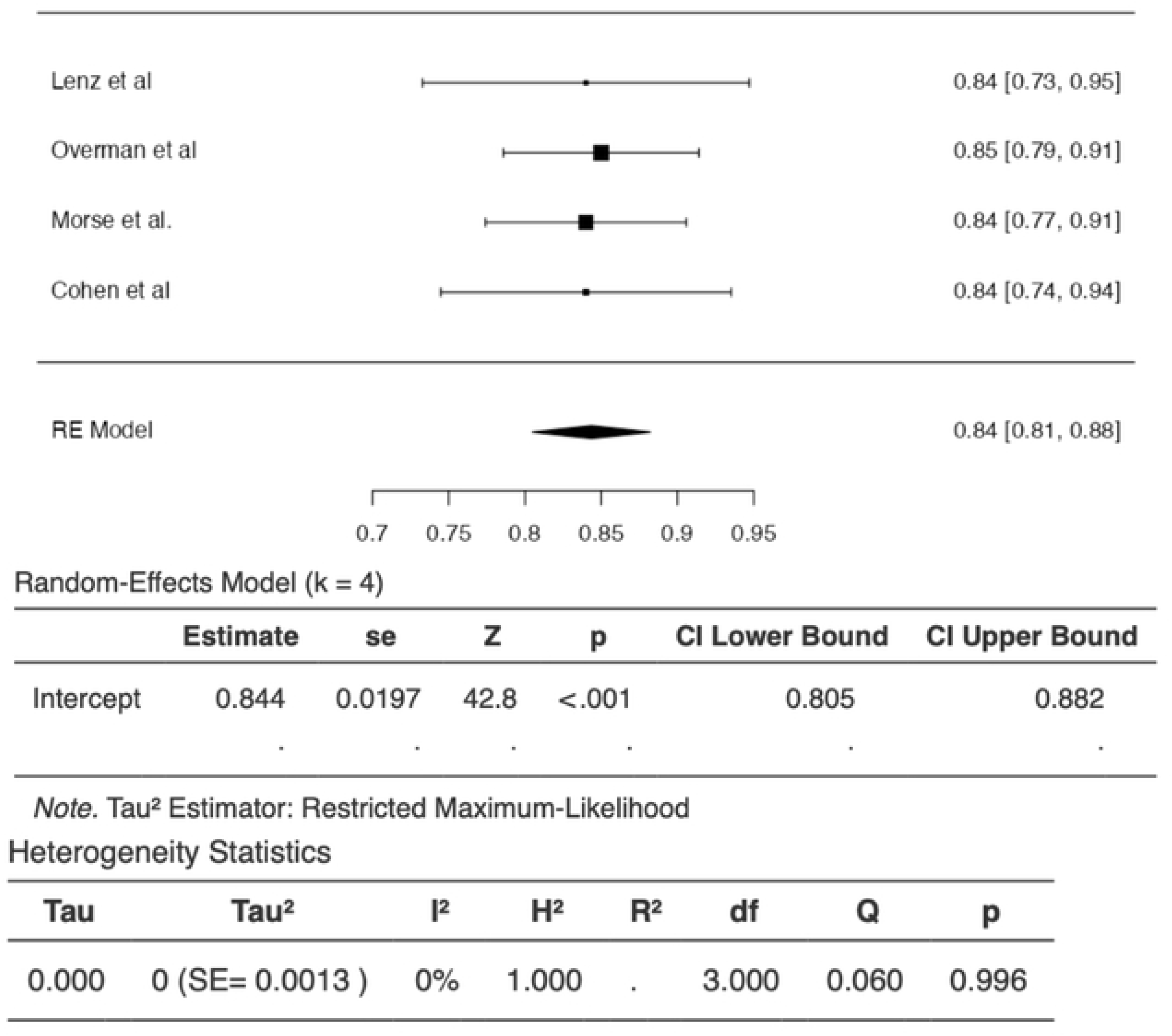
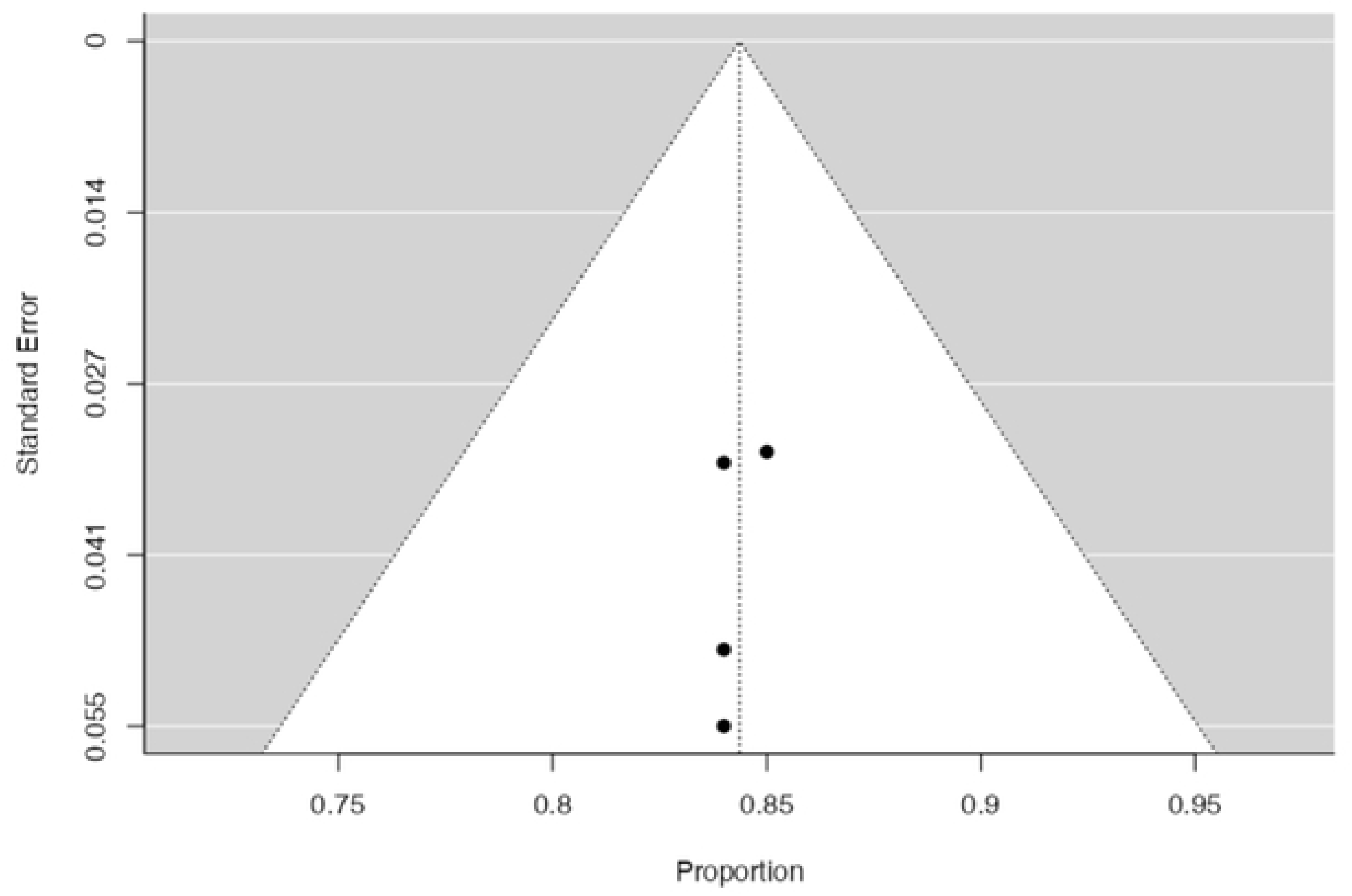
(A) Forest plot of OS Nivolumab and Ipilimumab intervention; (B) Funnel plot of OS nivolumab and Ipilimumab intervention.

Five studies that administered Pembrolizumab reported OS with an effect size of **0.56 [95% CI 0.37; 0.75, P<0.001]**. Heterogeneity was found to be high and the funnel plot shows evidence true of heterogeneity.

**Figure 8.**
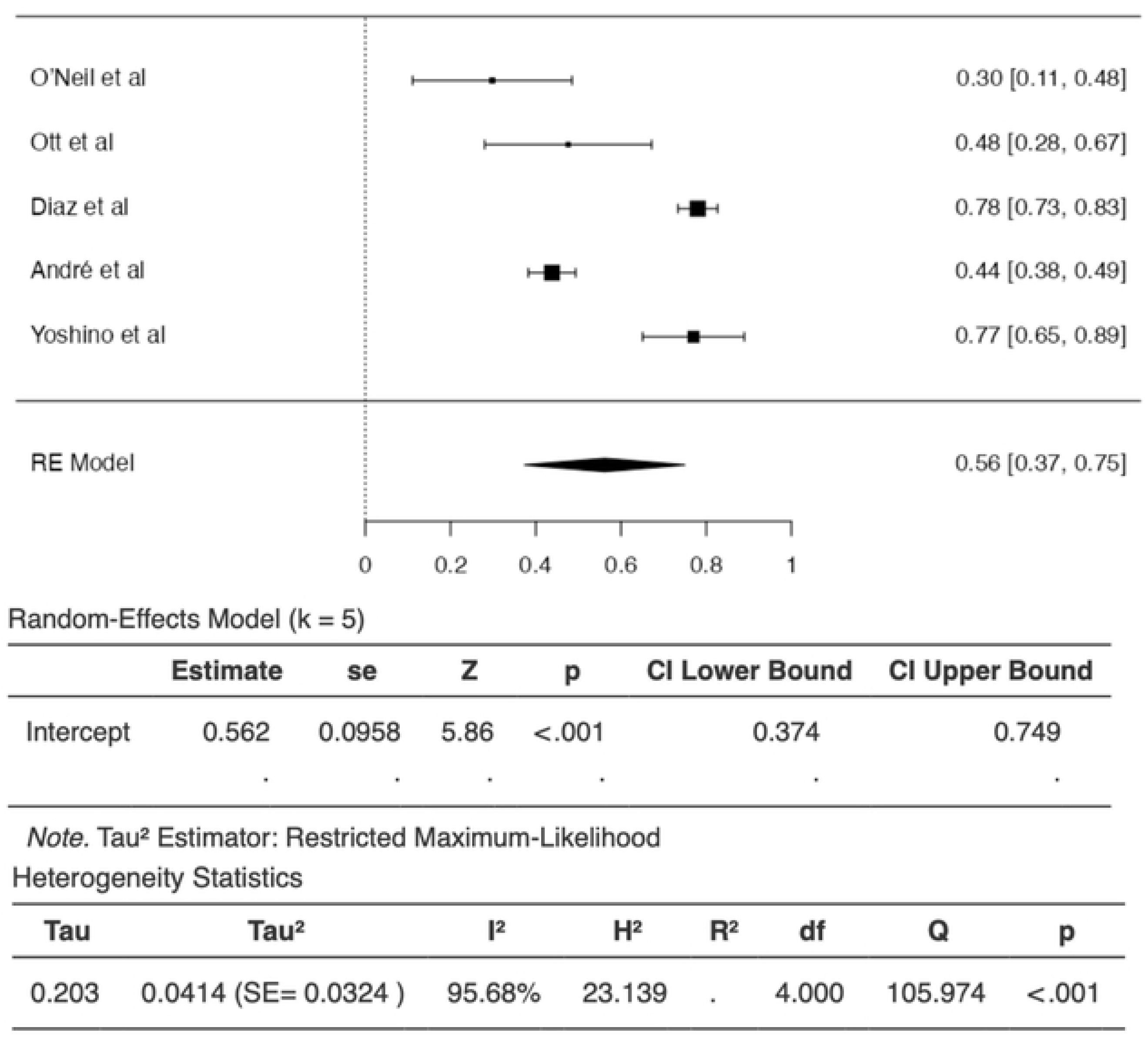
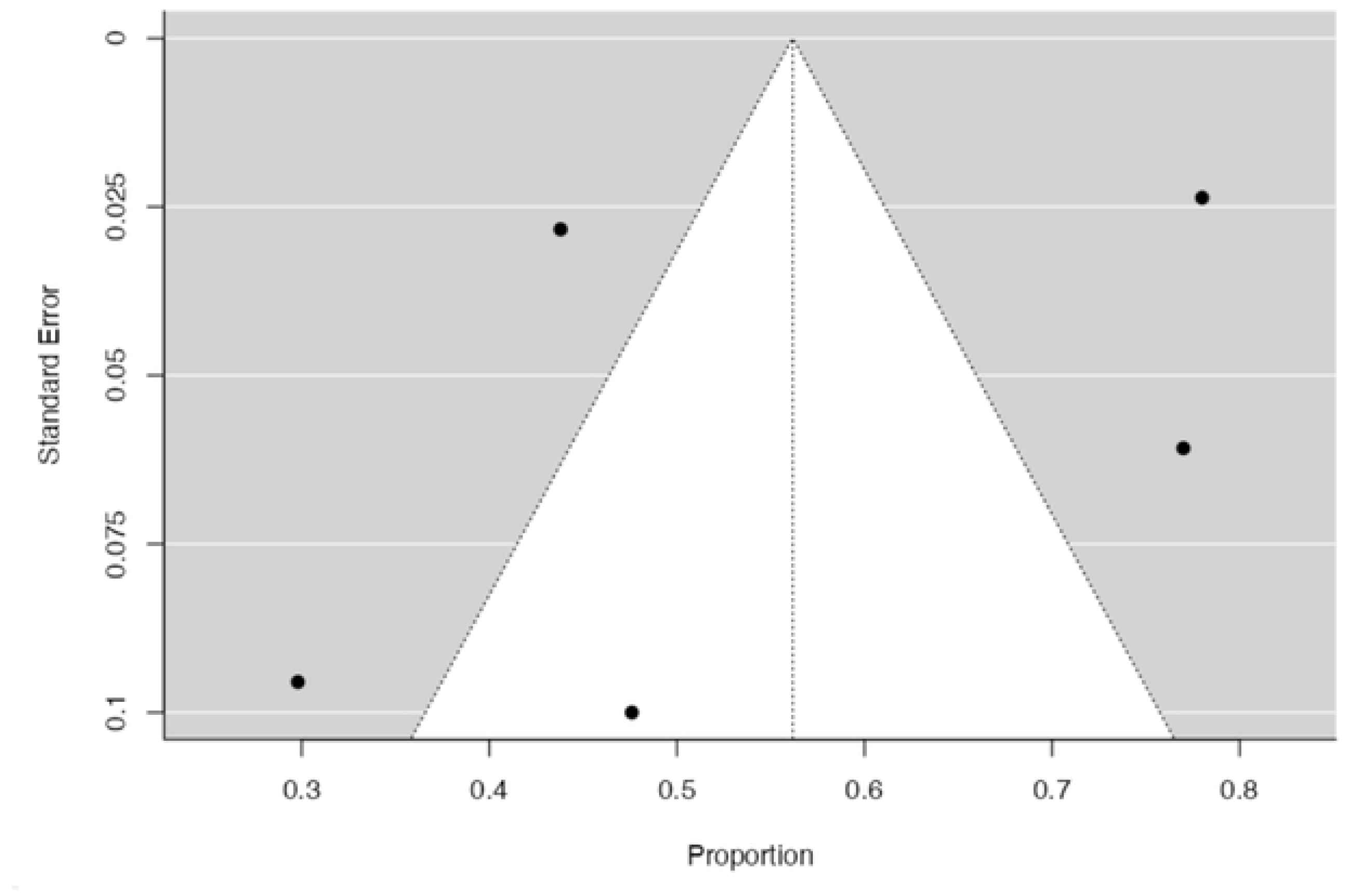
(A) Forest plot of OS Pembrolizumab intervention; (B) Funnel plot of OS Pembrolizumab intervention.

### Progression-free survival (PFS)

Three studies that administered Nivolumab reported PFS with an effect size of **0.53 [95% CI 0.43; 0.64, P<0.001]**. Heterogeneity was found to be low and the funnel plot shows no evidence true of heterogeneity

**Figure 9.**
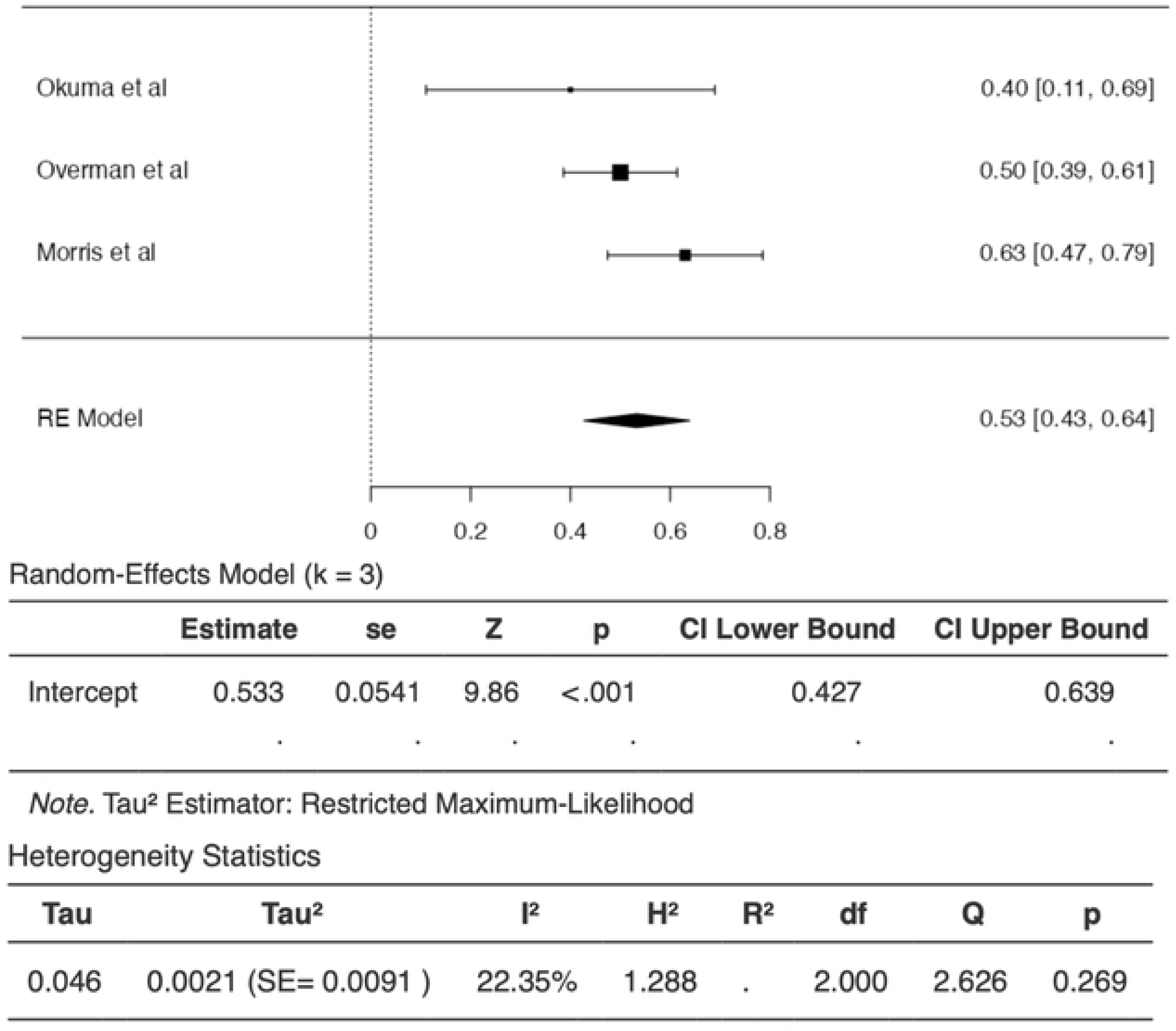
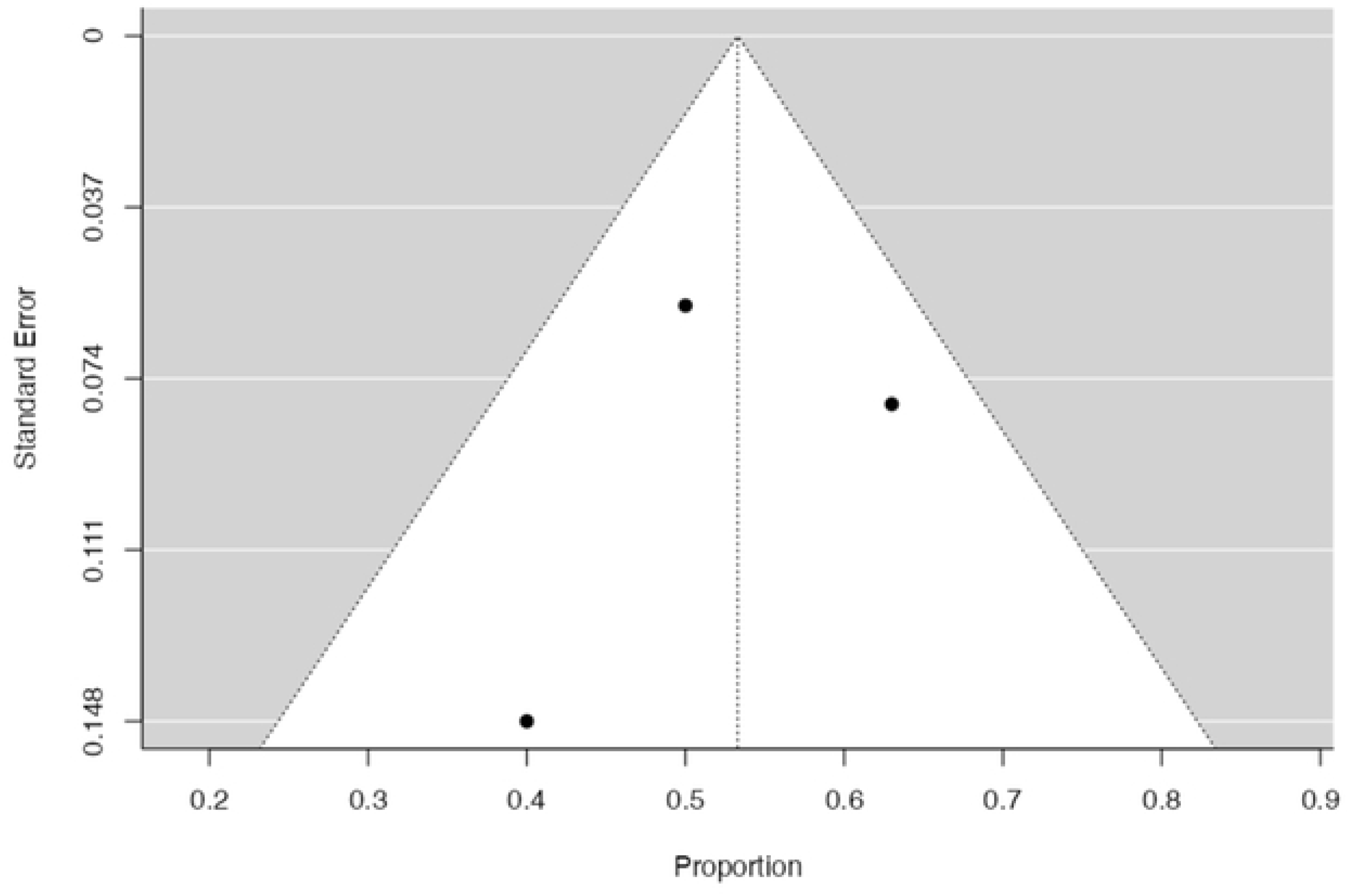
(A) Forest plot of PFS Nivolumab intervention; (B) Funnel plot of PFS Nivolumab intervention.

Four studies that administered Nivolumab and Ipilimumab reported PFS with an effect size of **0.72 [95% CI 0.67; 0.77, P<0.001]**. Heterogeneity was found to be absent and the funnel plot shows no evidence true of heterogeneity.

**Figure 10.**
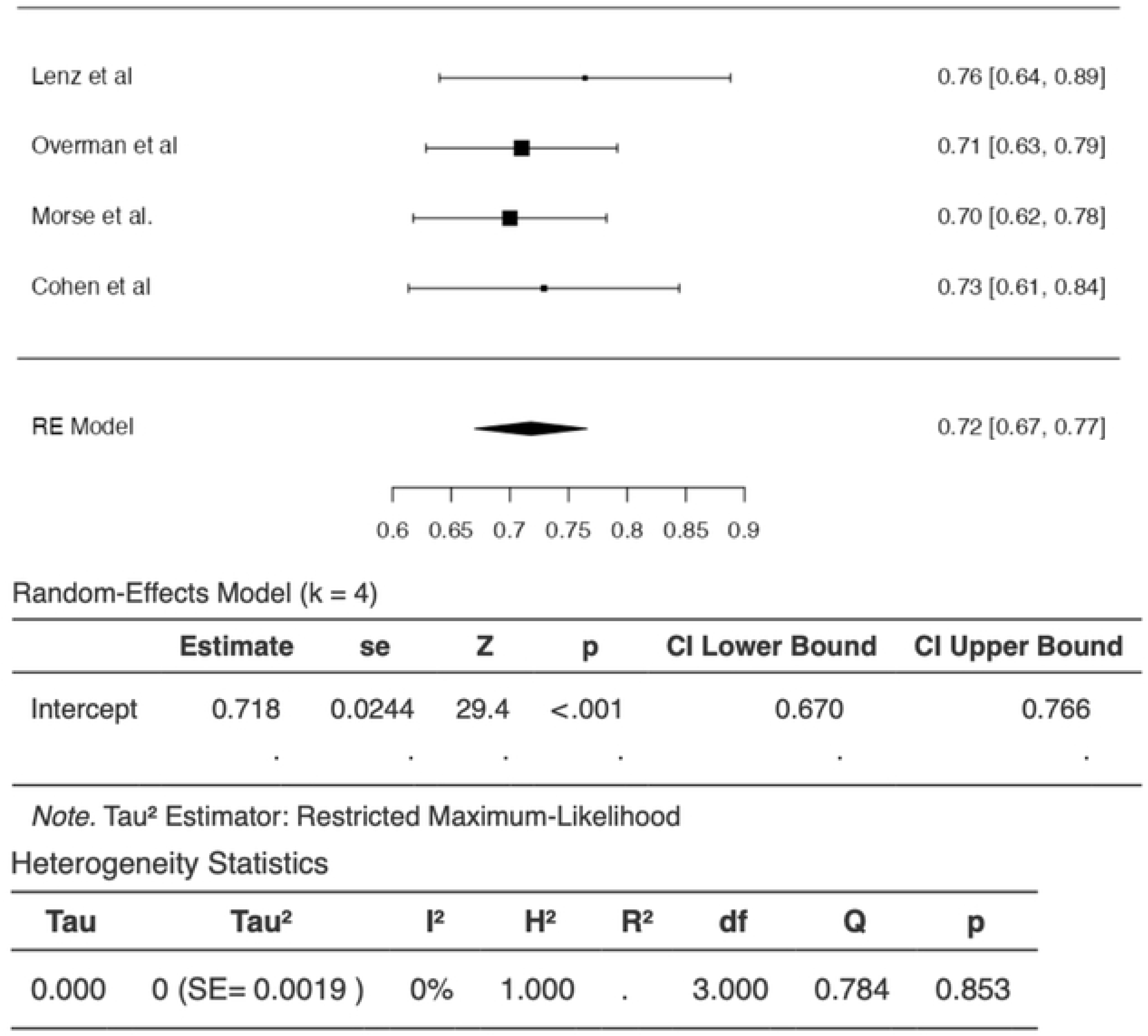
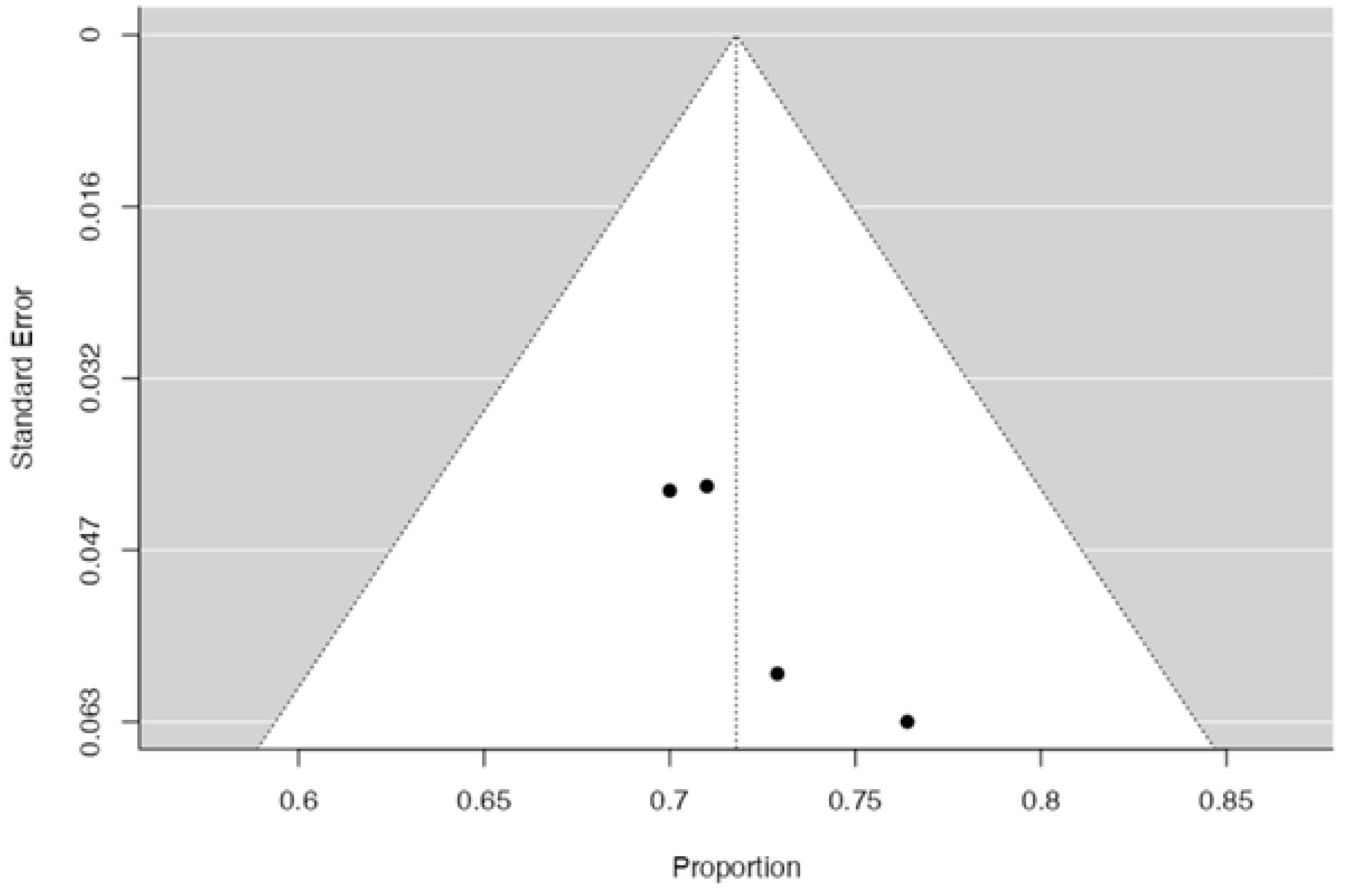
(A) Forest plot of OS Nivolumab and Ipilimumab intervention; (B) Funnel plot of OS Nivolumab and Ipilimumab intervention.

Five studies that administered Pembrolizumab reported PFS with an effect size of **0.53 [95% CI 0.34; 0.72, P<0.001]**. Heterogeneity was found to be high and the funnel plot shows evidence true of heterogeneity.

**Figure 11.**
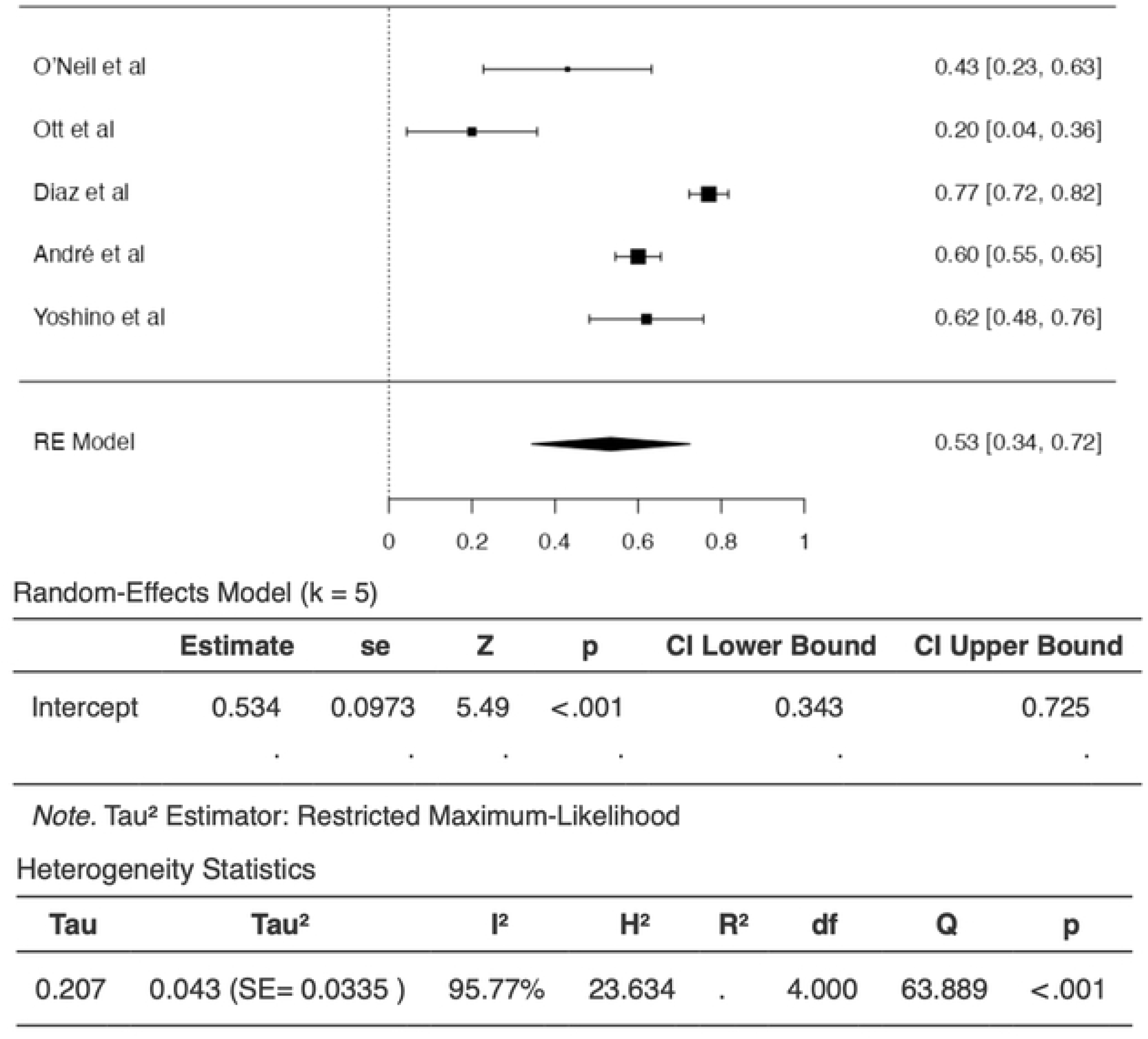
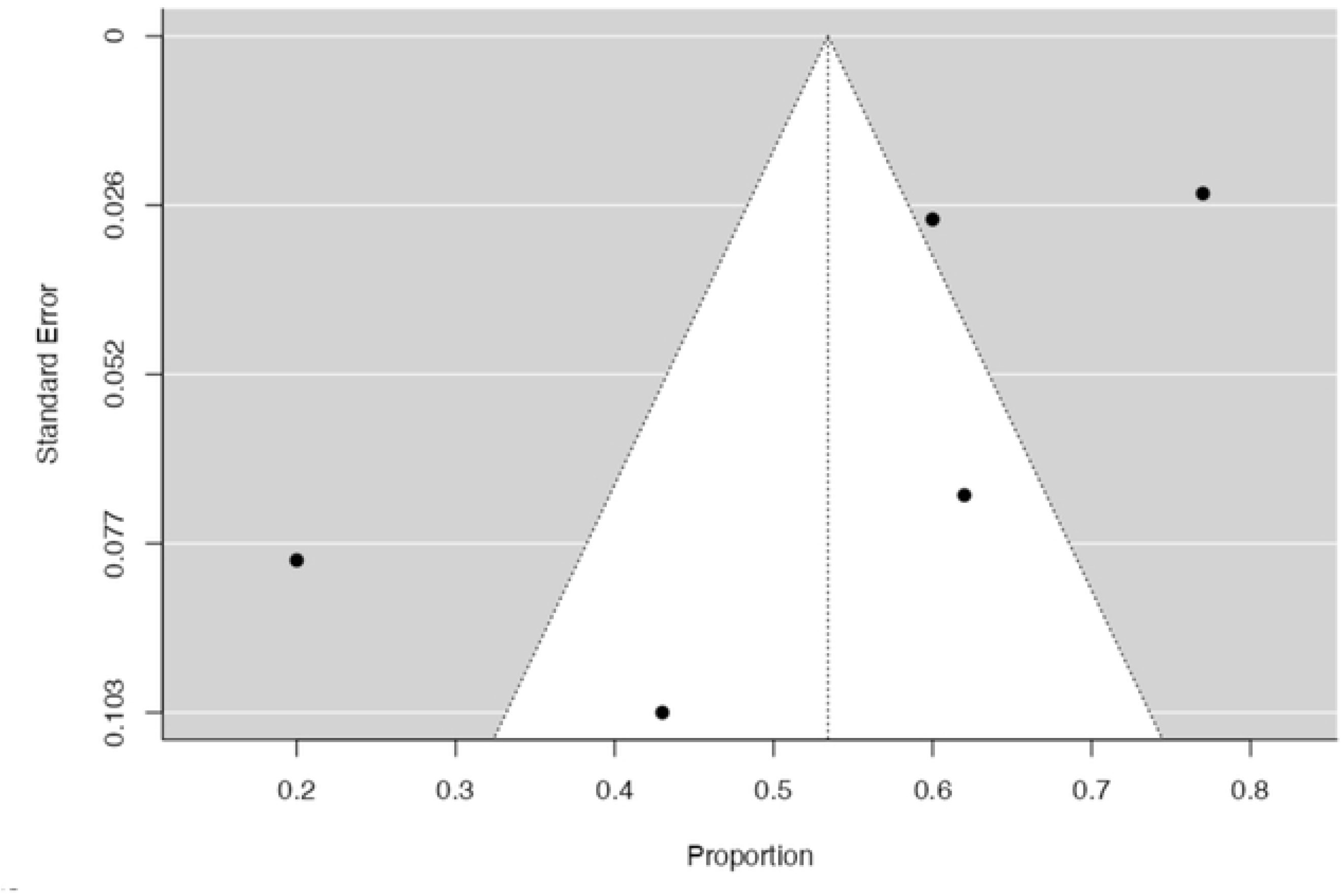
(A) Forest plot of PFS Pembrolizumab intervention; (B) Funnel plot of PFS Pembrolizumab intervention.

The effect size of each subgroup is summarized in Table 3 below.

**Table 3.** Size effects comparison of each subgroup.

**Objective Response Rate (ORR)**
| **Intervention** | **Effect Size (95% CI)** |
| --- | --- |
| Nivolumab | 0.34 (0.18; -0.49) |
| Nivolumab + Ipilimumab | 0.53 (0.42; -0.65) |
| Pembrolizumab | 0.34 (0.19; 0.48) |

**Overall Survival (OS)**
| **Intervention** | **Effect Size (95% CI)** |
| --- | --- |
| Nivolumab | 0.68 (0.52; 0.84) |
| Nivolumab + Ipilimumab | 0.84 (0.81; 0.88) |
| Pembrolizumab | 0.56 (0.37; 0.75) |

**Progression-Free Survival (PFS)**
| Intervention | Effect Size (95% CI) |
| --- | --- |
| Nivolumab | 0.53 (0.43; 0.64) |
| Nivolumab + Ipilimumab | 0.72 (0.67; 0.77) |
| Pembrolizumab | 0.53 (0.34; 0.72) |

## Discussion

Immunotherapy stands as a pivotal cornerstone in tumor control, and the exploration of combination immunotherapy involving multiple agents has emerged as a promising strategy. This comprehensive approach strategically intervenes in diverse immune response processes, encompassing chemoradiotherapy, targeted therapy, and interactions with immune pathways. By orchestrating a multifaceted anti-tumor immune response, combination immunotherapy seeks to mitigate the risk of drug resistance, marking it as a highly promising avenue for investigation[28].

Among the various combinations being studied, the concurrent administration of anti-PD-1 (nivolumab and ipilimumab) and anti-CTLA-4 (ipilimumab) has exhibited notable efficacy in tumor treatment[29,30]. This combination has demonstrated positive outcomes and substantial improvements in patients with metastatic melanoma, advanced renal cell carcinoma (RCC), and metastatic colorectal cancer (CRC)[30]. Cope et al. reported superior response rates in recurrent small cell lung cancer (SCLC) when treated with nivolumab plus ipilimumab, surpassing conventional chemotherapy and suggesting potential long-term survival benefits[31]. Additionally, Ready et al. provided evidence supporting the effectiveness and tolerability of the combination of nivolumab and low-dose ipilimumab as a first-line treatment for advanced/metastatic non-small cell lung cancer (NSCLC)[32].

In the realm of combination immunotherapy, the inclusion of pembrolizumab has further broadened the horizon of treatment modalities. Pembrolizumab, an anti-PD-1 agent, has demonstrated its efficacy in various malignancies[33,34]. The concurrent use of pembrolizumab with nivolumab and ipilimumab has shown promising outcomes, particularly in advanced RCC. Shao’s conditional survival analysis of first-line treatment highlighted that patients with advanced RCC undergoing pembrolizumab in combination with nivolumab and ipilimumab therapy exhibited a high survival rate compared to those receiving sunitinib[35]. This accumulating evidence underscores the potential synergistic effects of combining immunotherapeutic agents, notably anti-PD-1 and anti-CTLA-4, in conjunction with pembrolizumab, opening new avenues for the advancement of cancer treatment modalities.

The primary objective of this investigation was to assess the effectiveness and safety of immunotherapy drugs in the context of colorectal cancer. A comprehensive analysis was conducted, incorporating data from a total of 13 studies, encompassing a sample size of 1337 patients. The assessment of Efficacy (ORR, OS, PFS) for each groups with the investigation of adverse events and survival can validate the safety of this immunotherapy. However, AEs will only be covered descriptively in this study due to the different ways in which they have been presented in previous studies.

The combination of nivolumab and ipilimumab demonstrates the highest efficacy, with an ORR of 0.53, OS of 0.84, and PFS of 0.72. Pembrolizumab monotherapy also shows promising results, with an ORR of 0.34, OS of 0.56, and PFS of 0.53. However, the heterogeneity observed in some of the outcomes highlights the need for further studies to confirm these findings.

The combination of nivolumab and ipilimumab has been shown to be effective in various cancers, including colorectal carcinoma. The combination of these two medicine may enhance the immune response against cancer cells by targeting different checkpoints in the immune system. The high efficacy observed in our meta-analysis supports the use of this combination in mCRC patients[18].

Pembrolizumab, a PD-1 inhibitor, has been approved for the treatment of mCRC patients with dMMR or MSI-H tumors. The promising results observed in our meta-analysis suggest that pembrolizumab may be a viable treatment option for a broader range of mCRC patients. Further research is needed to confirm these findings and to explore the optimal treatment strategies for mCRC patients[19].

Ipilimumab+Nivolumab and Nivolumab monotherapy have been evaluated in earlier meta-analyses conducted across multiple publications with varying cancer types. It has been reported that the combination outperforms nivolumab monotherapy in terms of ORR and DCR, however patients may experience increased AEs[36]. One particular instance where using Ipilimumab+Nivolumab has been clearly demonstrated in multiple meta-analyses to result in improved ORR response[37–39]. Ipilimumab+Nivolumab use, however, still requires cautious consideration because of the possibility of excessive adverse effects. Therefore, more thorough research is required to determine how each type of cancer is affected by the combination of Ipilimumab and Nivolumab[40].

The Adverse Effects (AEs) found accoding to previous meta-analyses indicating a high incidence of AEs in the gastrointestinal system. However, in contrast, with the endocrine system, the use of anti-CTLA-4 or anti-PD-1 immunotherapy is reported to have a greater potential to cause endocrine issues (such as hypothyroidism, hyperthyroidism, thyroiditis, and adrenal insufficiency) in other cancers[41,42]. These findings are also in line with the study conducted by Almutairi AR, et al., 2020, who stated that Healthcare providers must remain vigilant in monitoring for immune-related adverse events (irAEs) subsequent to combined therapeutic interventions[43].

Additionally, particular attention should be directed towards gastrointestinal and cutaneous irAEs in the context of ipilimumab therapy. Furthermore, clinicians should be cognizant of the potential occurrence of irAEs associated with hyperglycemia, thyroid dysfunction, hepatic abnormalities, and musculoskeletal disorders following administration of nivolumab and pembrolizumab[43]. However, this treatment strategy is still preferable compared to conventional chemotherapy since it has better efficacy with the same adverse effects as stated by O’bryne K, et al., 2023, There was no statistically significant difference in the occurrence of adverse events (AEs) necessitating discontinuation between the combination of nivolumab and ipilimumab (NIVO + IPI) and pembrolizumab combined with chemotherapy, as well as between NIVO + IPI and pembrolizumab monotherapy. However, it is noteworthy that the incidence of treatment-related grade ≥3 AEs was higher with NIVO + IPI compared to pembrolizumab monotherapy[39].

### Strength and limitation

To our knowledge, this study represents the first systematic review and meta-analysis that compares both short-term and long-term effects of presently available anti-CTLA-4 and anti-PD-1 combination or monotherapies for treating patients with CRC. The PICOS criteria necessitated significant follow-up period in each trial to mitigate potential bias arising from heavily censored data in the tails of treatment arms during the observation period. Methodologically, the analysis was executed utilizing the meta library package in the Jamovi for the comprehensive analysis. The metabias package facilitated the execution of the Egger test, revealing evidence of heterogeneity for ORR, OS, and PFS outcomes. Additionally, the funnel plots also indicated evidence of heterogeneity for these outcomes. However, it is noteworthy that the prevalence of AEs, in general, remained at a low level, with instances of either absent or non-significant heterogeneity for certain AEs.

However, this study does have some limitations. Due to variations in the presentation of adverse events (AEs) across studies, with one individual potentially experiencing two or more AEs simultaneously, extracting data posed challenges for us. Additionally, It can be quite challenging to classify an AE as a potential immune-related AE (irAE), as this determination may vary among trials or observers. This variability could introduce detection bias, particularly because our analysis included both open-label and double-blind trials. Such variations might result in an overestimation of the incidence of potential irAEs. Nevertheless, we made efforts to mitigate this by only including patients treated with either anti-CTLA-4 or anti-PD-1 monotherapies or combination therapies, thereby excluding AEs that could be attributed to other treatments or immunotherapies. Furthermore, we employed the random-effects model for the meta-analyses and conducted subgroup analyses to address high heterogeneity, aiming to accommodate potential variations in the outcomes observed across the included studies. Moreover, significant statistical heterogeneity observed in most analyses is a crucial consideration. Variations in study designs, tumor stage, prior treatment history, and other characteristic among the included studies could have contributed to this heterogeneity.

## Conclusion

In conclusion, the exploration of combination immunotherapy involving anti-PD-1 and anti-CTLA-4 agents presents a promising avenue in cancer treatment, demonstrating notable efficacy across various cancers, including colorectal cancer (CRC). The concurrent use of nivolumab and ipilimumab, particularly, showcased superior efficacy in terms of objective response rate (ORR), overall survival (OS), and progression-free survival (PFS). Pembrolizumab monotherapy also exhibited promising results, highlighting its efficacy in treating CRC.

While the study, encompassing 13 studies with 1337 patients, provides valuable insights into the effectiveness and safety of immunotherapy in CRC, the identified heterogeneity in some outcomes emphasizes the necessity for additional research and exploration. Adverse effects, mainly observed in the gastrointestinal and endocrine systems, were generally low in incidence. Despite the higher incidence of treatment-related grade ≥3 adverse events with the nivolumab and ipilimumab combination, the superior efficacy observed supports its consideration as a viable treatment option. This study represents a pioneering systematic review comparing short-term and long-term effects of anti-CTLA-4 and anti-PD-1 therapies in CRC, acknowledging certain limitations and the need for ongoing investigation to refine and validate these findings.

## Data Availability

All relevant data are within the manuscript and its Supporting Information files

## Data availability

### Underlying data

All data underlying the results are available as part of the article and no additional source data are required.

### Reporting guidelines

Mendeley Data doi: 10.17632/zhvz39sggy.1.

Data are available under the terms of the Creative Commons Attribution 4.0 International license (CC-BY 4.0).

## Funding

The author(s) declared no funding information available.

## Competing interests

No competing interests were disclosed.

## Grant information

The author(s) declared that no grants were involved in supporting this work.

## Acknowledgment

We would like to express our sincere gratitude to the Indonesian Health Research Foundation for their invaluable contribution in reviewing and enhancing the quality of this systematic review and meta-analysis. Their expertise and insightful feedback have significantly enriched the rigor and comprehensiveness of our research. We acknowledge the Foundation’s commitment to advancing healthcare research and their dedication to fostering excellence in scientific inquiry. This collaboration has been instrumental in shaping the robustness of our study, and we extend our heartfelt appreciation for their support in promoting the advancement of health knowledge.

## Notes

### Competing Interest Statement

The authors have declared no competing interest.

### Funding Statement

The author(s) received no specific funding for this work.

